# VEXAS syndrome is characterized by blood and tissues inflammasome pathway activation and monocyte dysregulation

**DOI:** 10.1101/2022.10.12.22281005

**Authors:** Olivier Kosmider, Céline Possémé, Marie Templé, Aurélien Corneau, Francesco Carbone, Eugénie Duroyon, Twinu-Wilson Chirayath, Marine Luka, Camille Gobeaux, Estibaliz Lazaro, Roderau Outh, Guillaume Le Guenno, François Lifermann, Marie Berleur, Chloé Friedrich, Cédric Lenormand, Thierry Weitten, Vivien Guillotin, Barbara Burroni, Pierre Sohier, Jay Boussier, Lise Willems, Selim Aractingi, Léa Dionet, Pierre-Louis Tharaux, Béatrice Vergier, Pierre Raynaud, Hang-Korng Ea, Mickael Ménager, Darragh Duffy, Benjamin Terrier

## Abstract

Acquired mutations in the *UBA1* gene, occurring in myeloid cells and resulting in expression of a catalytically impaired isoform of the enzyme E1, were recently identified in patients with severe adult-onset auto-inflammatory syndrome called VEXAS (vacuoles, E1 enzyme, X-linked, autoinflammatory, somatic). The precise physiological and clinical impact of these mutations remains poorly defined.

Here, we studied a unique prospective cohort of individuals with severe autoinflammatory disease with (VEXAS) or without (VEXAS-like) *UBA1* somatic mutations and compared with low-risk myelodysplastic syndromes (MDS) and aged gender-matched healthy controls. We performed an integrated immune analysis including multiparameter phenotyping of peripheral blood leukocytes, cytokines profiling, bulk and single-cell gene expression analyses and skin tissue imaging mass cytometry.

Focusing on myeloid cells, we show that monocytes from *UBA1*-mutated individuals were quantitatively and qualitatively impaired and displayed features of exhaustion with aberrant expression of chemokine receptors. Within affected tissues, pathological skin biopsies from VEXAS patients showed an abundant enrichment of CD16^+^ CD163^+^ monocytes adjacent to blood vessels and M1 macrophages, possibly promoting local inflammation in part through STAT3 activation. In peripheral blood from VEXAS patients, we identified a significant increase in circulating levels of many proinflammatory cytokines, including IL-1β and IL-18 which reflect inflammasome activation and markers of myeloid cells dysregulation. Gene expression analysis of whole blood confirmed the role of circulating cells in the IL-1β and IL-18 dysregulation in VEXAS patients and revealed a significant enrichment of TNF-α and NFκB signaling pathways that could mediate cell death and inflammation. Single-cell analysis confirmed the inflammatory state of monocytes from VEXAS patients and allowed us to identify specific molecular pathways that could explain monocytopenia, especially the activation of PANoptosis and a deficiency in the TYROBP/DAP12 axis and β-catenin signaling pathway. Together, these findings on monocytes from patients with *UBA1* mutations provide important insights into the molecular mechanisms involving the mature myeloid commitment in VEXAS syndrome and suggest that the control of the undescribed inflammasome activation and PANoptosis could be novel therapeutic targets in this condition.

**GRAPHICAL ABSTRACT:** 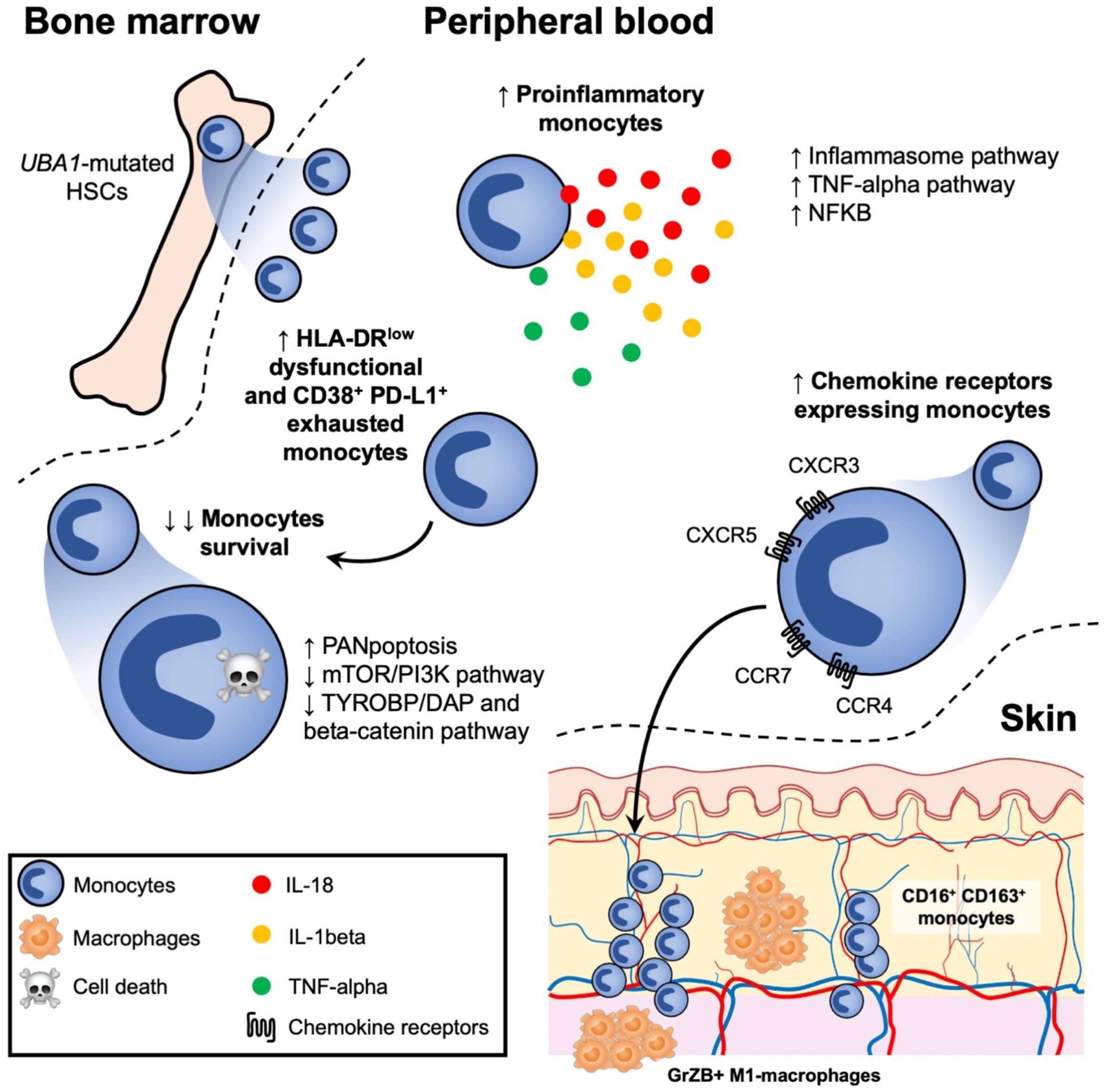

## INTRODUCTION

Somatic missense mutations affecting methionine-41 (p.Met41) or splice site in the *UBA1* gene, which result in the expression of a catalytically impaired isoform of the protein, were recently identified in patients with severe adult-onset autoinflammatory disease (Beck et al., 2020; Templé et al., 2021). The E1 ubiquitin-activating enzyme UBA1 is responsible for initiation of the activation, conjugation and ligation of ubiquitin to target protein substrates, playing a critical role in regulating protein homeostasis and cellular processes (Groen and Gillingwater, 2015). Using a genotype-driven approach from the analysis of peripheral blood exome sequence data, Beck et al. identified a disorder named VEXAS (vacuoles, E1 enzyme, X-linked, autoinflammatory, somatic) syndrome (Beck et al., 2020; Templé et al., 2021). Prevalence of VEXAS syndrome is not yet known but the disease frequency is likely underestimated. VEXAS syndrome was initially exclusively described in males showing somatic mutations in *UBA1* (Beck et al., 2020). However, we and others described VEXAS syndrome in female patients, consequent to acquired somatic mutation and X monosomy in hematological cells (Arlet et al., 2021). Clinical manifestations include fever, neutrophilic cutaneous, arthralgias, pulmonary inflammation, chondritis, and vasculitis, supporting the recruitment and accumulation of inflammatory cells in these tissues (Beck et al., 2020; Ferrada et al., 2021; Grayson et al., 2021). Additionally, patients with VEXAS suffer from a spectrum of hematologic manifestations mainly including macrocytic anemia, thrombocytopenia, thromboembolic disease, and progressive bone marrow failure, sharing some features with myelodysplastic syndrome (MDS) (Grayson et al., 2021). Vacuoles are localized predominantly in promyelocytes, myelocytes and erythroid precursors in the bone marrow from VEXAS patients, but what these vacuoles contain is not yet clear (Grayson et al., 2021). VEXAS syndrome is commonly refractory to conventional disease-modifying antirheumatic drugs and biological targeted therapies may show partial efficacy, suggesting a highly dysregulated inflammatory response. The hypomethylating agent azacytidine, that can be effective in MDS associated or not with inflammatory diseases, showed some efficacy in VEXAS patients (Comont et al., 2022), as well as ruxolitinib, a potent and selective oral inhibitor of both JAK1 and JAK2 protein kinases (Bourbon et al., 2021). So far, the only curative option in VEXAS syndrome with severe manifestations was reported to be allogeneic stem cell transplantation (Diarra et al., 2022).

Somatic mutations affecting the p.Met41 were described in hematopoietic stem cells and peripheral blood myeloid cells but not in mature lymphocytes nor fibroblasts. Mutant cells showed decreased ubiquitylation activating cellular stress responses that lead to upregulation of the unfolded-protein response (UPR), dysregulation of autophagy, and a shared gene expression signature consistent with the activation of multiple innate immune pathways (Beck et al., 2020). However, little is known about the immunological features and the molecular mechanisms induced by this impaired ubiquitylation.

To explore the inflammatory mechanisms induced by *UBA1* somatic mutations and to identify therapeutic targets, we used an integrative approach based on clinical and biological data, in-depth phenotypical analysis of whole blood immune cells, cytokine profiling, whole blood bulk RNA analysis, single-cell RNA sequencing of peripheral blood mononuclear cells (PBMCs), and tissue imaging by mass cytometry on skin biopsies, on a large group of *UBA1*-mutated individuals with autoinflammatory disease (VEXAS) in comparison to autoinflammatory individuals without *UBA1* mutation (VEXAS-like), low risk MDS without features of inflammation and aged gender-matched healthy individuals. We demonstrate that circulating monocytes from *UBA1*-mutated individuals, when compared with VEXAS-like, MDS and healthy controls, were quantitatively and qualitatively impaired and displayed features of exhaustion associated with aberrant expression of chemokine receptors, and dysregulation of IL-1β and IL-18 processing consistent with activation of the inflammasome pathway. Within affected tissues bearing *UBA1* somatic mutations, an abundant enrichment of CD16^+^ CD163^+^ monocytes and M1 macrophages was present, possibly promoting local inflammation in part through STAT3 activation. Transcriptomic analyses also revealed a significant enrichment of TNF-α and NFκB signaling that could mediate cell death and inflammation. We identified at the single cell level molecular pathways involved in monocyte dysregulation, especially a defective TYROBP/DAP12 and β-catenin signaling pathway and the activation of the proinflammatory programmed cell death PANoptosis.

## RESULTS

### Characteristics of the cohort

We analyzed the immune response elicited by *UBA1* somatic mutations was analyzed in a unique cohort of individuals with severe autoinflammatory diseases showing *UBA1* mutation (VEXAS) (n=38), nonmutated aged gender-matched individuals with severe autoinflammatory diseases (VEXAS-like) (n=26), MDS (n=4) and aged gender-matched used as healthy controls (n=12) (**Fig. 1a and Supplementary Table 1**). In all cases, fresh material was used to analyze *UBA1* mutations and to generate multi-Omics data (**Fig. 1b)**.

**Fig. 1.**
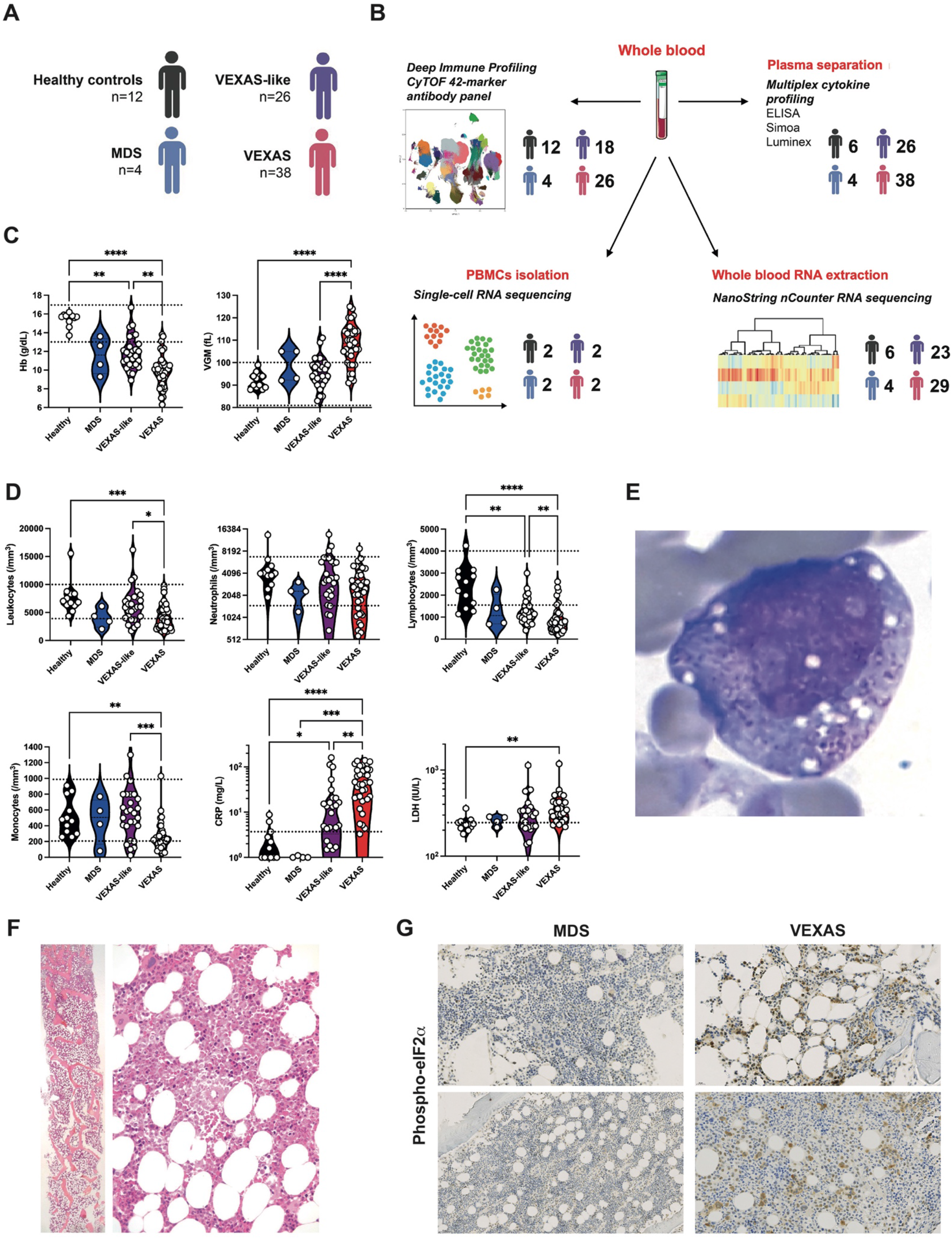
Clinical and laboratory findings in patients with VEXAS syndrome, VEXAS-like, MDS and healthy controls. (**A, B**) 38 individuals with VEXAS syndrome, 26 individuals with VEXAS-like, 4 with MDS and 12 aged gender-matched healthy controls were included, and samples were assessed using deep immune profiling, plasma multiplex cytokine profiling, whole blood RNA extraction and single-cell RNA sequencing. (**C**) Hemoglobin (Hb) and mean corpuscular volume (MCV) from patients with VEXAS syndrome, VEXAS-like, MDS and healthy controls. Each dot represents a single patient. (**D**) Leukocytes, neutrophils, lymphocytes and monocytes count, C-reactive protein (CRP) and lactate dehydrogenase (LDH) from patients with VEXAS, VEXAS-like, MDS and healthy controls. Each dot represents a single patient. (**E**) Bone marrow aspiration showing vacuoles restricted to myeloid and erythroid precursor cells in a VEXAS patient. (**F**) Bone marrow biopsy from a VEXAS patient showing overrepresentation of the myeloid cell lineage with mature and immature forms. (**G**) Immunohistochemical staining of phosphorylated eIF2α on bone marrow biopsy from MDS and VEXAS patients, showing expression of p-EIF2α only in VEXAS. P values were determined by the Kruskal-Wallis test, followed by Dunn’s post test for multiple group comparisons. *P <0.05; **P <0.01; ***P <0.001, ****P <0.0001.

All individuals were male with a median age of 74 (55-88) years for VEXAS, 70 (54-90) years for VEXAS-like, 76 (70-88) years for MDS and 60 (55-85) years for healthy controls at time of sampling. Each of the VEXAS individuals had either one of the three somatic variants of codon 41 in *UBA1* (p.Met41Thr in 17, p.Met41Val in 9,or p.Met41Leu in 6) or a splice mutation affecting the p.Met41 in 6 cases. The majority of VEXAS individuals had recurrent fevers, arthralgias/arthritis, neutrophilic dermatosis and cutaneous vasculitis, pulmonary involvement (**Supplementary Table 1**), macrocytic anemia (**Fig. 1c**), leukopenia, mainly lymphocytopenia and monocytopenia, and increased C-reactive protein and lactate dehydrogenase levels (**Fig. 1d**), and bone marrow vacuolization restricted to myeloid and erythroid precursor cells (**Fig. 1e**). Bone marrow biopsies from VEXAS patients (n=5), compared with VEXAS-like (n=2) and MDS (n=3), showed heterogeneity in morphology and cellularity but predominantly overrepresentation of the myeloid cell lineage with many mature and immature forms, and slight abnormalities of erythroblasts and megakaryocytes (example from VEXAS patient shown in **Fig. 1f**). Myeloid cells in bone marrow from VEXAS patients only, but not in VEXAS-like nor MDS patients, showed expression of phosphorylated eIF2α (p-EIF2α) confirming the activation of cellular stress responses leading to upregulation of the UPR and dysregulation of autophagy as described before (Beck et al., 2020) (**Fig. 1g**).

### An aberrant phenotype of circulating monocytes in *UBA1*-mutated individuals

We first performed a multiparameter phenotyping of peripheral blood leukocytes in all individuals using mass cytometry (**Supplementary Table 2**). Immunoprofiling of peripheral blood from VEXAS patients revealed a dramatic decrease of circulating monocytes, primarily involving intermediate (CD14^+^ CD16^+^) and nonclassical (CD14^lo^ CD16^+^) monocytes (**Fig. 2a**). By integrating data from patients with VEXAS, VEXAS-like, MDS and healthy controls and subjecting them to dimensionality reduction using non-supervised Uniform Manifold Approximation and Projection (UMAP) (Becht et al., 2018) (**Fig. 2b and 2c**), we analyzed monocyte subsets from all patient groups. Analysis revealed dramatic differences in the repartition of monocyte subsets in VEXAS in comparison with other groups (**Fig. 2d**). Volcan plots of differentially represented monocyte clusters showed an increased proportion of clusters 8, 11, 14, 15 and 20 in VEXAS compared to healthy controls, and a decreased proportion of clusters 5, 6, 7, 13 and 16 (**Fig. 2e and Extended Data Fig. 1**). Heatmap representing the relative expression of cell surface markers identified clusters 8, 11, 14 and 15 as dysfunctional HLA-DR^lo^ classical monocytes, and cluster 20 as exhausted HLA-DR^lo^ CD38^+^ PD-L1^hi^ nonclassical monocytes (**Fig. 2f and Extended Data Fig. 2a**). Cell surface expression of HLA-DR on monocytes, evaluated by mean metal intensity (MMI), was also significantly decreased in VEXAS patients (**Extended Data Fig. 2b**). Clusters 5, 6, 7, 13 and 16, significantly decreased in VEXAS, identified in contrast functional HLA-DR^hi^ classical, intermediate and nonclassical monocytes (**Fig. 2f**). Similarly, compared to VEXAS-like, VEXAS patients had an increased proportion of the same dysfunctional classical and nonclassical monocytes and exhausted HLA-DR^lo^ CD38^+^ PD-L1^hi^ nonclassical monocytes, while the same functional HLA-DR^hi^ monocytes were decreased (**Fig. 2e and 2f**). Consistent with these findings, whole blood expression of activation-related genes (Palojärvi et al., 2013)—such as *HLADRB1* and *CD86*—were significantly decreased, and expression of exhaustion-related genes (Pradhan et al., 2021)—such as *PLAUR and RELB*—were significantly increased in VEXAS patients compared to other groups (**Extended Data Fig. 2c**). VEXAS syndrome has been associated with various clinical manifestations, related to massive influx of innate immune cells. We analyzed the expression of chemokines receptors on monocytes. Dysfunctional and exhausted monocytes from VEXAS (clusters 14 and 20) showed significantly higher expression of CXCR3, CXCR5, CCR4 and CCR7 compared to VEXAS-like, MDS and healthy controls (**Fig. 2g and Extended Data Fig. 2d**). These cells expressed low levels of CD14 supporting that they were nonclassical monocytes. Interestingly, CXCR3 expression is able to promote transendothelial migration of cells to sites of inflammation, through the interaction of CXCR3 with CXCL9, 10, and 11. CXCR3 and CCR4 were also shown to be expressed by immune cells in inflamed skin tissue, both receptors having been associated with dermal recruitment of immune cells, an important finding given the high frequency of skin lesions in VEXAS (Al-Banna et al., 2014). Also, CXCR5, a receptor for CXCL13, and CCR7, a receptor for CCL19 and CCL21, are key regulators of cell localization in secondary lymphoid organs (Ohl et al., 2003), which could explain monocytes migration to lymph nodes and spleen. Monocyte chemotactic factor chemokine (C-C motif) ligand 2 (CCL2), also called MCP-1, was increased only in the blood of VEXAS patients, whereas transcripts of its receptor *CCR2* were dramatically decreased (**Extended Data Fig. 2e**). Such discrepancy between CCR2 expression and CCL2/MCP-1 protein levels was previously shown to represent a feedback mechanism in the regulation of the chemotactic response of monocytes/macrophages (Fantuzzi et al., 1999).

**Fig. 2.**
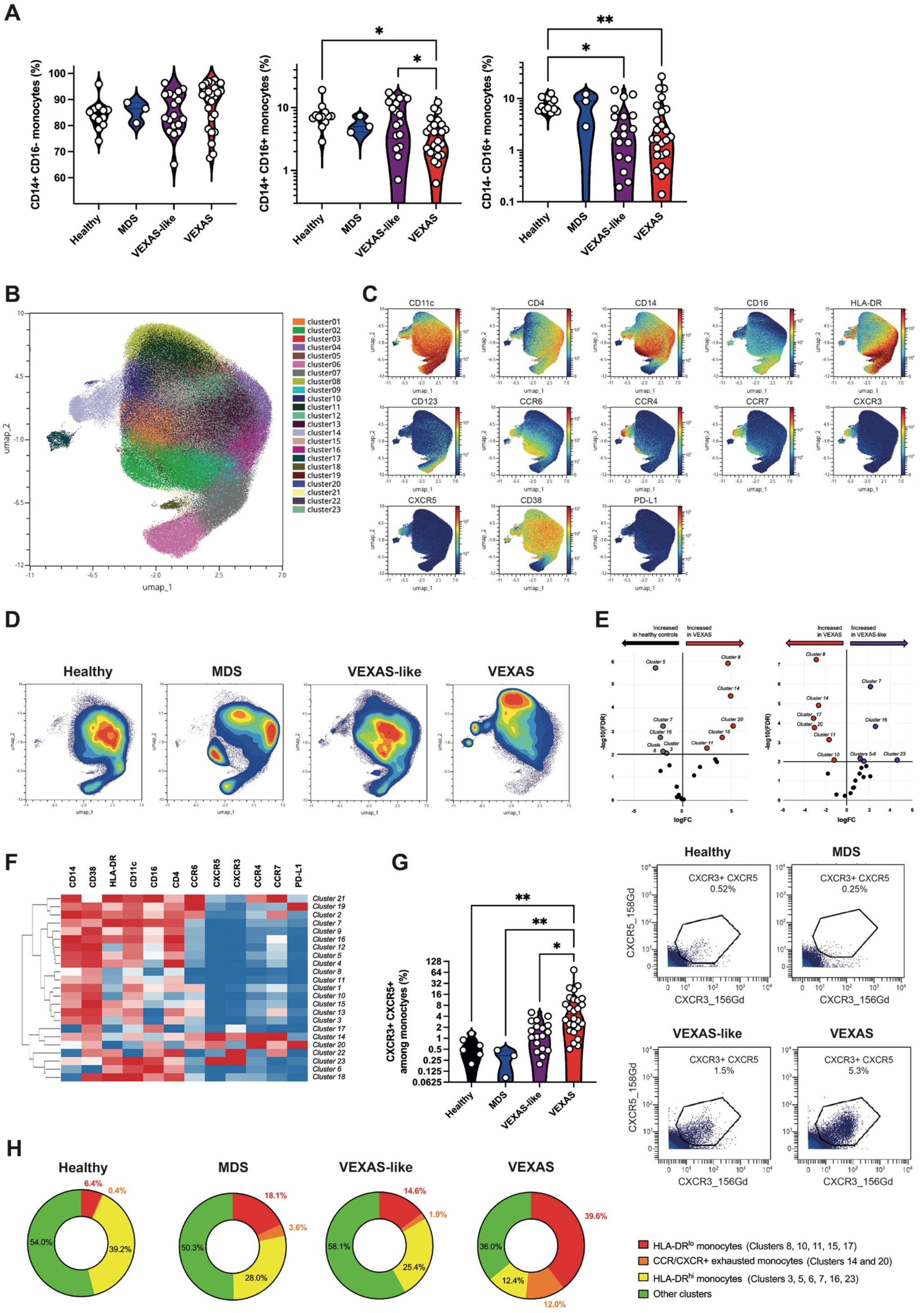
Phenotyping of peripheral blood monocytes in VEXAS syndrome. (**A**) Proportions (frequencies) of classical (CD14^+^, CD16!), intermediate (CD14^+^, CD16^+^), and nonclassical (CD14^lo^, CD16^+^) monocytes among blood monocytes from VEXAS patients, VEXAS-like patients, MDS and healthy controls, were analyzed. Each dot represents a single patient. (**B**) Non-supervised Uniform Manifold Approximation and Projection (UMAP) of blood monocytes. Cells are automatically separated into spatially distinct subsets according to the combination of markers that they express. (**C**) UMAP of blood monocytes stained with 13 markers and measured with mass cytometry. (**D**) UMAP colored according to cell density across patients’ groups. Red indicates the highest density of cells. (**E**) Volcan plots of differentially represented monocyte subsets showing clusters increased or decreased in VEXAS patients compared to healthy controls, and VEXAS patients compared to VEXAS-like. (**F**) Heatmap representation of all monocyte clusters, ordered by hierarchical clustering and expression of the cell surface markers. (**G**) Proportion (frequencies) of CXCR3+ CXCR5+ expressing monocytes. Each dot represents a single patient. Gating strategy for the analysis of expression of CXCR3 and CXCR5 on blood monocytes, Illustrative dot plots are shown and enumeration of percentages of CXCR3+ CXCR5+ cells. (**H**) Pie chart showing the proportion of HLA-DR^lo^ dysfunctional monocytes, exhausted monocytes expressing chemokine receptors, HLA-DR^hi^ functional monocytes, and other clusters not significantly different between groups. Numbers on the pie charts indicate the median proportion of each monocytes subsets in each group. P values were determined by the Kruskal-Wallis test, followed by Dunn’s post test for multiple group comparisons. *P <0.05; **P <0.01.

Altogether, these data suggest that changes in the abundance and phenotype of monocyte subsets within the peripheral blood cell population, with dramatically reduced monocytes enriched in dysfunctional and exhausted subsets showing elevated chemokine receptors expression (**Fig. 2h**), characterize VEXAS patients. Dramatic decrease in circulating monocytes could suggest either an increased cell death and/or an enhanced monocyte migration into inflamed tissues.

### Enrichment of CD16^+^CD163^+^ monocytes and M1 macrophages in skin lesions from VEXAS patients

Formalin-fixed paraffin-embedded (FFPE) slides from skin biopsy specimens from 3 VEXAS at time of active lesions were analyzed (**Fig. 3a**). As previously reported (Zakine et al., 2021), we detected by Sanger sequencing on both paired bone marrow or peripheral blood and skin biopsy specimens from VEXAS patients the same *UBA1* mutation (data not shown). Immunohistochemical staining showed abundant monocytes expressing CD68 and MPO within the dermis of inflamed skin lesions, whereas neutrophils expressing CD15 were less numerous (**Fig. 3b**). As in bone marrow samples, mononuclear cells in the skin showed increased expression of p-EIF2α (**Fig. 3c**).

**Fig. 3.**
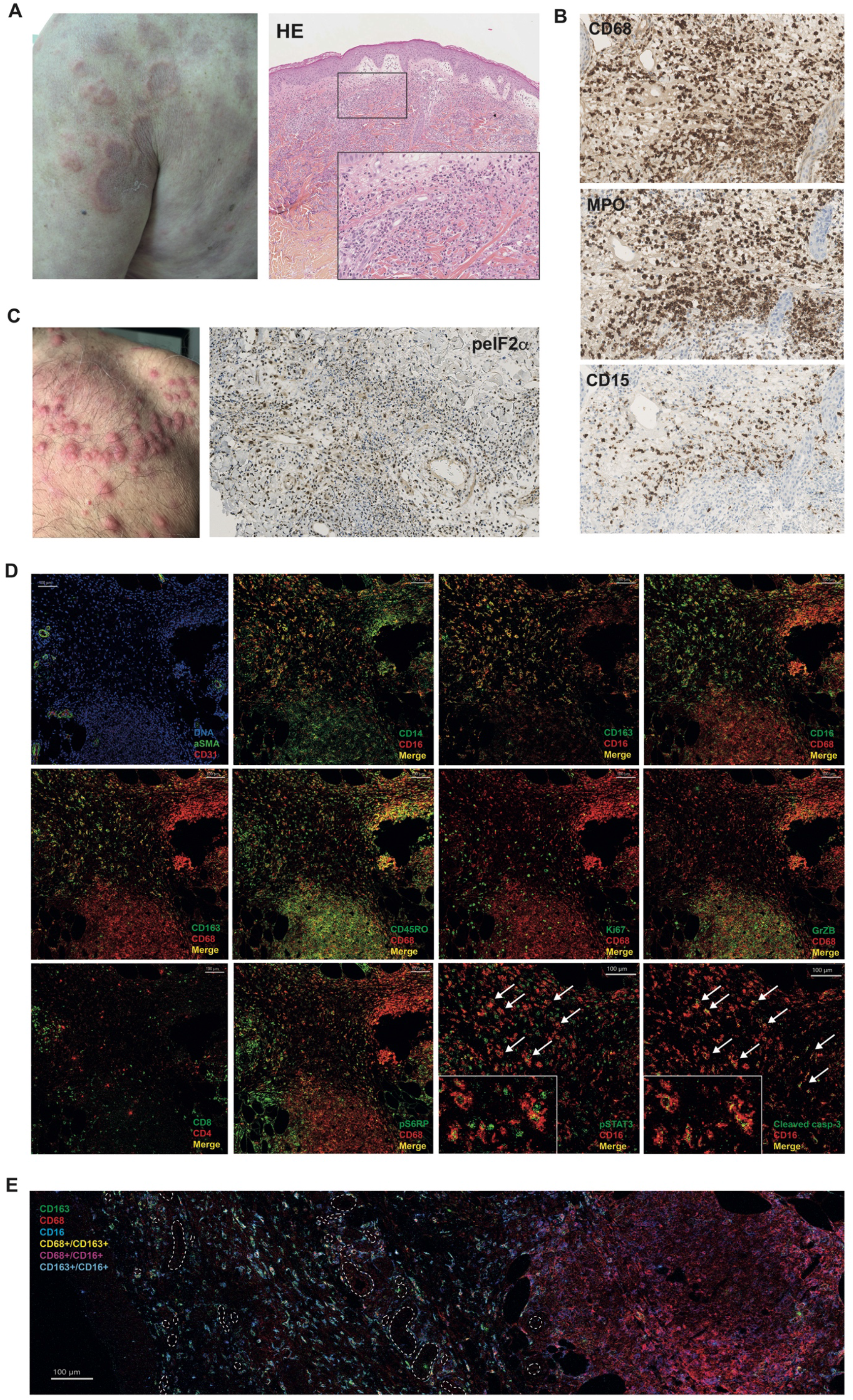
Characterization and anatomical localization of myeloid cells within skin lesion from VEXAS patients. (**A**) Picture of neutrophilic dermatosis and hematoxylin-eosin staining of skin biopsy of a VEXAS patient. Illustrative picture is shown. (**B**) Immunohistochemical staining of CD68, myeloperoxidase (MPO) and CD15 of the same skin biopsy. Illustrative picture is shown. (**C**) Picture of neutrophilic dermatosis of another VEXAS patient and immunohistochemical staining of phosphorylated eIF2α on skin biopsy from a VEXAS patient, showing expression of p-EIF2α in skin lesions. Illustrative picture is shown. (**D**) Imaging mass cytometry on skin biopsy showing abundant nonclassical (CD14^lo^CD16^+^) and intermediate (CD14^+^CD16^+^) monocytes expressing CD163, adjacent to blood vessels, some of them showing colocalization with phospho-STAT3 and cleaved caspase-3 (white arrows), and CD68^+^CD163^-^ M1 macrophages forming clusters away from blood vessels. Illustrative pictures are shown. (**E**) Imaging mass cytometry on skin biopsy from another VEXAS patient showing abundant nonclassical (CD14^lo^CD16^+^) and intermediate (CD14^+^CD16^+^) monocytes expressing CD163, and CD68^+^CD163^-^ M1 macrophages forming clusters. Blood vessels are represented by the dotted line.

We then used imaging mass cytometry to determine anatomical localization of inflammatory cells within target tissues from VEXAS patients, using a combination of markers (**Extended Data Fig. 3**). Focusing on monocytes/macrophages, nonclassical (CD14^lo^ CD16^+^) and intermediate (CD14^+^ CD16^+^) monocytes expressing CD163 were abundant in skin lesions from VEXAS cases (**Fig. 3d**), adjacent to blood vessels, suggesting trafficking from the periphery to areas of inflammation. Such CD16^+^ CD163^+^ monocyte population was described in necrotizing enterocolitis in premature infants as playing an important role in the inflammatory process and trafficking to sites of inflammation, suggesting that these cells could be one of the pathogenic drivers of inflammation in target tissues (Olaloye et al., 2021). Some CD16^+^ monocytes showed colocalization with phospho-STAT3 as well as many surrounding cells, supporting the proinflammatory phenotype of infiltrating cells (**Fig. 3d**). STAT3 is a critical signaling molecule activated by a variety of extracellular stimuli resulting in phosphorylation on tyrosine residues. Persistently activated STAT3 in monocytes was demonstrated to promote cell survival and invasion in cancer models and also to promote inflammation, especially through the NFκB and IL-6/JAK pathways (Yu et al., 2009). Such CD16^+^ monocytes were also shown to secrete proinflammatory cytokines (Narasimhan et al., 2019). These data reinforce the hypothesis that CD16^+^ CD163^+^ inflammatory monocytes may play diverse functions supporting inflammation in skin lesions from VEXAS patients. Additionally, few CD16^+^ monocytes showed colocalization with cleaved caspase-3, supporting that they were undergoing an active cell death process (**Fig. 3d**).

Next to CD16^+^ CD163^+^ monocyte-enriched areas, we identified CD68^+^ CD163^-^ macrophages forming clusters or aggregates further away from blood vessels (**Fig. 3e**). The scavenger receptor CD163 is a highly specific marker for the M2 subpopulation. Accumulation of CD68^+^ CD163^-^ macrophages within skin biopsies suggests abundant proinflammatory M1 macrophages next to CD16^+^ CD163^+^ monocytes. M1 macrophages are usually induced by LPS and IFN-γ and activate chemotaxis and cell migration functions (Orecchioni et al., 2020). CD68^+^ CD163^-^ macrophages from skin lesions also expressed granzyme B, suggesting that granzyme B may play a role in macrophage functions and pathogenicity associated with skin lesions.

Overall, these results support a scenario in which *UBA1*-mutated inflammatory monocytes aberrantly expressing chemokine receptors could be attracted into target tissues and partly rescued from apoptosis, promoting local inflammation mediated in part through STAT3 activation.

### VEXAS patients display an increase in proinflammatory cytokines consistent with inflammasome activation

To further characterize inflammatory responses implicated in VEXAS, we analyzed plasma samples from patients with VEXAS, VEXAS-like, MDS and healthy controls by Luminex Multiplex assays, conventional and digital ELISA to quantify a total of 52 soluble mediators. Unsupervised principal components analysis (PCA) separated patients with VEXAS syndrome from the other groups on dimension 2, driven by inflammatory cytokines (IL-6, IL-18), IL-1 receptor antagonist (IL-1RA) and myelomonocytic markers (calprotectin and galectin-3) (**Fig. 4a**).

**Fig. 4.**
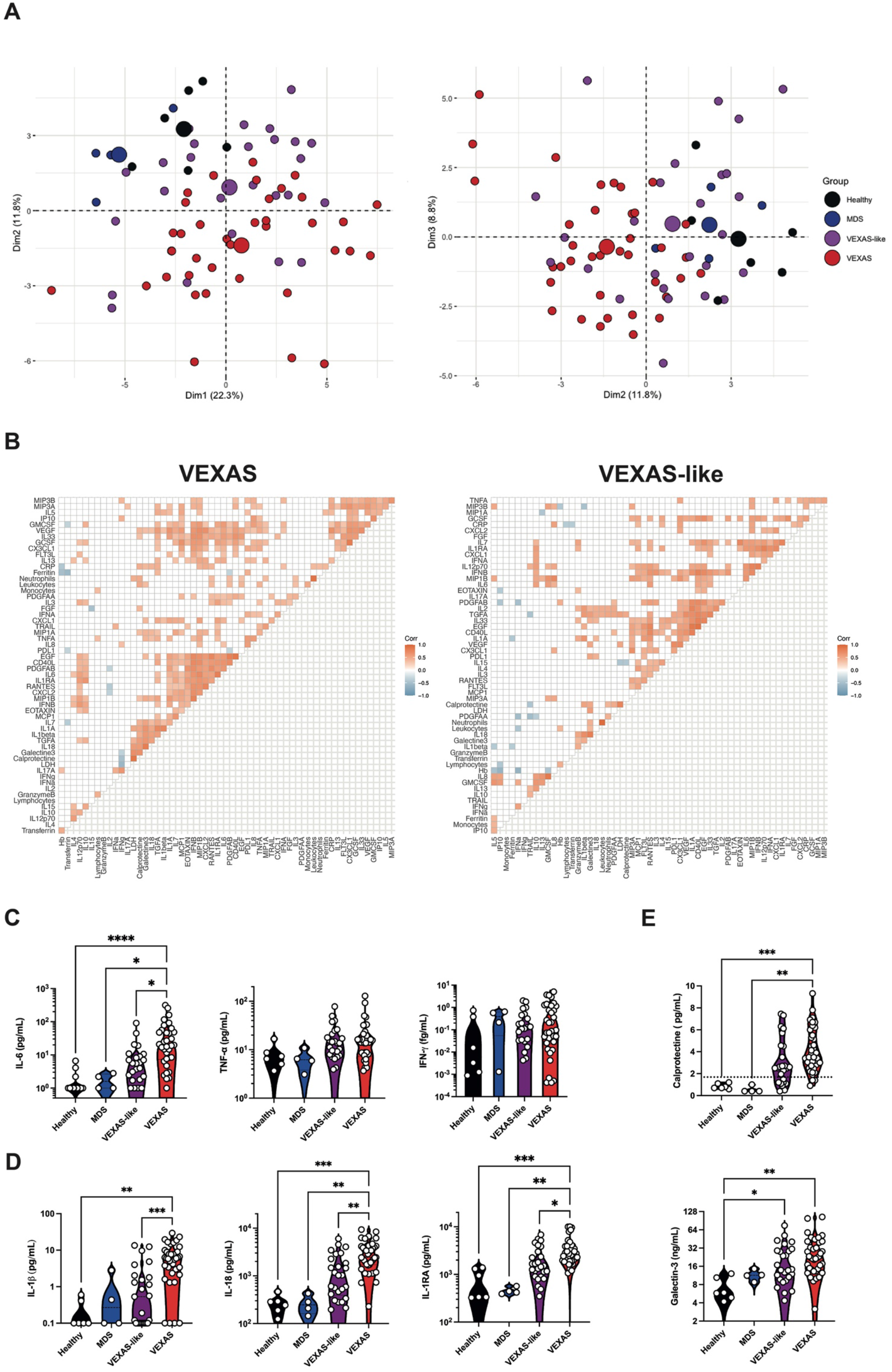
Inflammatory cytokine and chemokines profiling of VEXAS syndrome. (**A**) Principal component analysis of the cytokine and chemokine data according to dimensions 1, 2 and 3. Median values for each patients’ group are plotted with the large colored circle. Each dot represents a single patient. (**B**) Correlation matrices across all values of cytokines and chemokines from patient blood, comparing patients with VEXAS and VEXAS-like. Only significant correlations (<0.05) are represented as squares. Pearson’s correlation coefficients from comparisons of cytokine and chemokine measurements within the same patients are visualized by color intensity. (**C**) IL-6, TNF-α and IFN-γ from patients with VEXAS, VEXAS-like, MDS and healthy controls. Each dot represents a single patient. (**D**) IL-1β, IL-18 and IL-1RA from patients with VEXAS, VEXAS-like, MDS and healthy controls. Each dot represents a single patient. (**E**) Calprotectin and galectin-3 from patients with VEXAS, VEXAS-like, MDS and healthy controls. Each dot represents a single patient. P values were determined by the Kruskal-Wallis test, followed by Dunn’s post test for multiple group comparisons. *P <0.05; **P <0.01; ***P <0.001, ****P <0.0001.

We next tried to correlate the measurements of these soluble proteins (**Fig. 4b**). We observed a ‘core VEXAS signature’ that was defined by the following inflammatory mediators, which correlated positively with each other: IL-1α, IL-1β, IL-18, TGF-α, IL-7, galectin-3 and calprotectin. We also observed in VEXAS two additional clusters: the first defined by inflammatory cytokines—IL-6, IL-1RA—and proinflammatory chemokines—CCL2/MCP-1, eotaxin, CCL4/MIP-1β, CXCL2, RANTES, PDGF-AB and EGF—and the second defined mainly by proinflammatory chemokines and growth factors—CCL20/MIP-3α, CCL19/MIP-3β, CX3CL1, IL-10, G-CSF, GM-CSF and VEGF. VEXAS-like patients mainly showed a more heterogeneous signature defined by IL-2, IL-33 and TGF-α, chemokines and growth factors. As suggested by PCA and correlation matrix, prominent inflammatory cytokines included IL-6, a key player of exacerbated inflammatory responses, that was dramatically increased in VEXAS compared to other groups, whereas tumor necrosis factor–α (TNF-α), another key driver of inflammation, and IFN-γ levels were not significantly elevated (**Fig. 4c**). IL-1β and IL-18, potent proinflammatory cytokines for which maturation and secretion is governed by the inflammasome (Chan and Schroder, 2020), were also dramatically increased in VEXAS patients (**Fig. 4d**). These findings corroborated with the detection of high amounts of circulating IL-1RA, indicating an active antagonism of IL-1 in VEXAS patients (**Fig. 4d**).

Calprotectin and galectin-3 were specifically increased in the peripheral blood of VEXAS patients compared to other groups, highly supporting dysregulation of the myelomonocytic compartment (**Fig. 4e**). S100A8/S100A9 alarmin calprotectin is known to be released under inflammatory conditions by myeloid cells and promotes NFκB activation (Riva et al., 2012) and secretion of multiple inflammatory proteins, such as IL-6 (Wang et al., 2018). Galectin-3 (formerly known as Mac-2), is secreted from monocytes/macrophages (Cherayil et al., 1989; Satoh et al., 2012) and regulate monocyte/macrophage adhesion, chemotaxis, and apoptosis (Sano et al., 2000).

These data highlight broad inflammatory changes, especially involving concomitant release of IL-1β and IL-18 governed by the inflammasome and markers of myeloid cells dysregulation in VEXAS patients.

### Proinflammatory transcriptional signatures characterize VEXAS patients

To investigate the immunological transcriptional signatures that characterize VEXAS patients, we quantified the expression of 594 immune-related genes in peripheral blood cells by using the Nanostring Human Immunology v2 panel kit. Supervised hierarchical clustering showed differences in gene expression between aged gender-matched healthy controls and MDS in one hand, and VEXAS and VEXAS-like cases on the other hand (**Fig. 5a**). Unsupervised principal components analysis (PCA) also showed distinct clustering of VEXAS syndrome from the other groups (**Fig. 5b**).

**Fig. 5.**
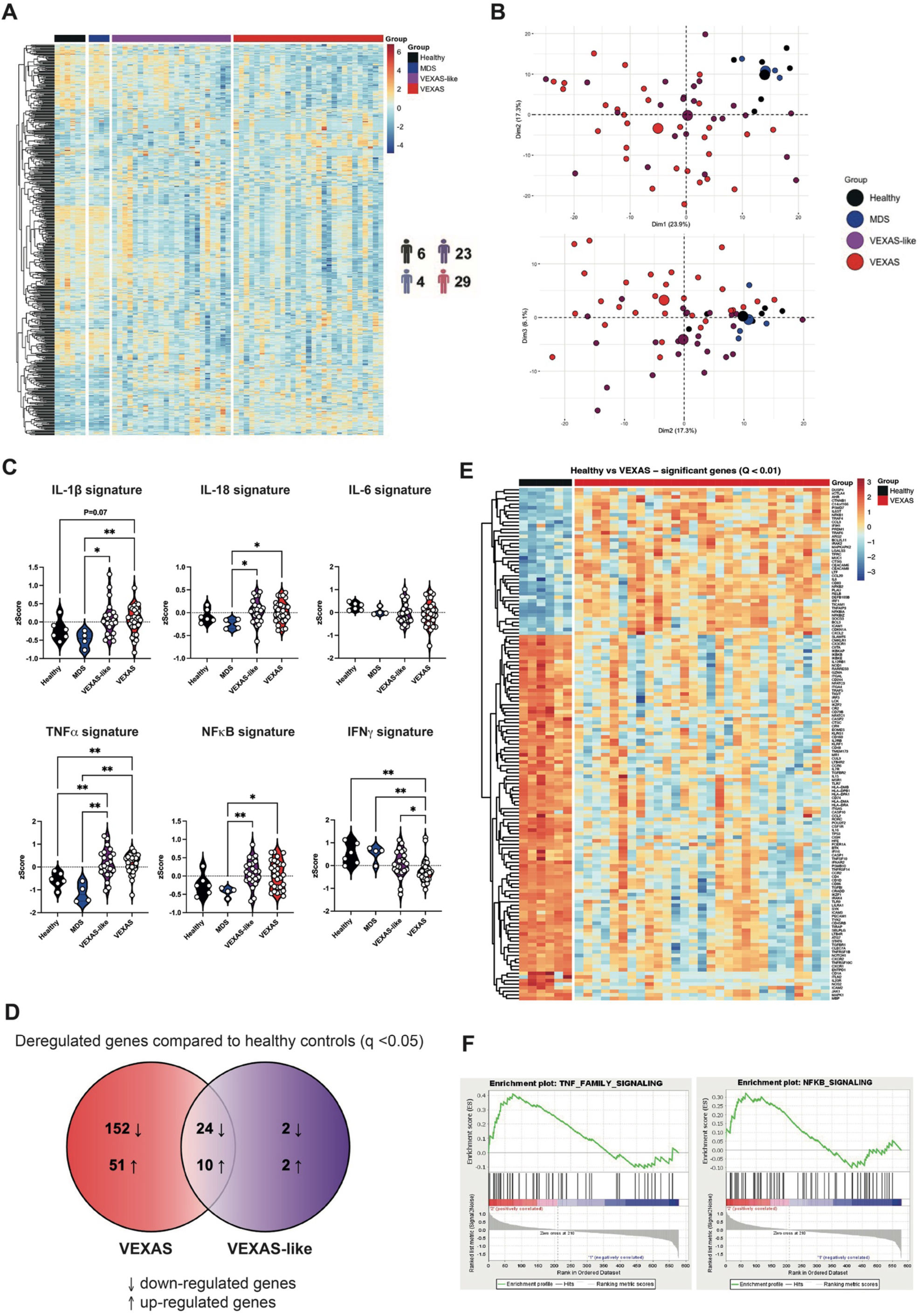
Immunological transcriptional signatures in VEXAS syndrome. (**A**) Heatmap representation of 579 immunological genes measured by Nanostring approach, ordered by hierarchical clustering in healthy controls (n=6), MDS (n=4), VEXAS-like (n=23), and VEXAS (n=29). (**B**) Principal component analysis of the transcriptional data according to the 4 groups of patients. (**C**) Comparison of IL-1β, IL-18, IL-6, TNF-α, NFκB and type II IFN gene signatures expression in healthy controls, MDS, VEXAS-like and VEXAS patients. Each dot represents a single patient. (**D**) Venn diagram representation showing significant differential gene expression in VEXAS or VEXAS-like patients versus healthy controls. (**E**) Heatmap representation of differentially expressed genes (q-value <0.01) in healthy controls (n=6) and VEXAS (n=29) patients, ordered by hierarchical clustering. (**F**) Gene set enrichment analysis of pathways enriched in VEXAS versus healthy controls, i.e. TNF-α signaling and NFκB signaling. P values were determined by the Kruskal-Wallis test, followed by Dunn’s post test for multiple group comparisons. *P <0.05; **P <0.01.

Guided by previous results from plasma cytokine measurement (**Fig. 4c and 4d**), we used specific lists of genes to calculate zScore for each transcriptional signature, i.e. type II IFN, TNF-α, IL-1β, IL-18, IL-6 and NFκB (type II IFN, TNF-α, and IL-1β signatures were based on previous data from Urrutia et al.(Urrutia et al., 2016), while IL-18, IL-6 and NFκB signatures were based on the Nanostring Immunology panel annotation file or other databases informations, **Supplementary Table 3**). IL-1β, IL-18, TNF-α and NFκB signatures were upregulated in both VEXAS and VEXAS-like patients, whereas the IL-6 signature was not significantly different between groups and the type II IFN (IFN-γ) signature was significantly reduced in VEXAS cases compared to other groups (**Fig. 5c**).

We then used Venn diagram representation to visualize distinction between gene expression data from VEXAS, VEXAS-like and healthy controls (MDS patients were not included in this analysis due to too few patients). We compared the list of significantly dysregulated genes (Mann-Whitney test, q-value <0.05) in the respective VEXAS and VEXAS-like analyses versus healthy controls (**Fig. 5d**). Among the 237 genes dysregulated in VEXAS compared to controls, 202 were specifically deregulated in VEXAS of which 152 were down-regulated and 51 up-regulated. The 10 most down-regulated genes were *CCR2, CSF1R, CMKLR1, CIITA, MSR1, CX3CR1, CASP10, CD86, CISH and TLR7*, while the 10 most up-regulated genes were *PLAU, NFKBIA, LTF, TNFAIP3, CTSG, CEACAM6, CXCL2, CEACAM8, CD83 and CCL20* (**Supplementary Table 4**). Notably, 34 of the 237 genes differentially dysregulated between VEXAS and controls were also dysregulated in the VEXAS-like versus controls (**Supplementary Table 5**), while only 4 genes were specifically dysregulated in VEXAS-like confirming the greater heterogeneity of VEXAS-like patients.

Heatmap representation of supervised hierarchical clustering showed differences in gene expression between healthy controls and VEXAS patients (**Fig. 5e**). Gene set enrichment analysis of pathways enriched between healthy controls and VEXAS showed that genes encoding proteins involved in TNF-α pathway signaling and NFκB signaling were upregulated in VEXAS (**Fig. 5f, Extended Data Fig. 4a and 4b**), while those encoding proteins in adaptive immunity, MHC class II antigen presentation and phagocytosis degradation were downregulated (gene set enrichment analysis enrichment score with *q* value <0.2).

Finally, we analyzed Nanostring data using Ingenuity Pathway Analysis (Qiagen) software to study connections between differentially regulated genes, upstream regulators and signaling pathways potentially responsible for the observed differences in gene expression. Increased expression of *CTNNB1, CXCL8, IFNB1, NFKB1* and *TNF* in VEXAS cases pointed to the signaling pathway integrating Fas-Associated protein with Death Domain (FADD) as a major upstream regulator with a zScore of 2.2, the latter being involved into the ripoptosome pathway with caspase-8 (*CASP8*) and *RIPK1* (**Extended Data Fig. 4c**).

Overall, our data suggest that VEXAS patients exhibit an upregulation of IL-1β and IL-18 signatures at the transcriptional level suggesting inflammasome activation, and TNF-α family and NFκB signaling that could mediate cell death and inflammation.

### Single-cell RNA sequencing of PBMCs highlights dysregulated proinflammatory and cell death signatures in monocytes from VEXAS syndrome

To further characterize the features of monocyte populations that characterize VEXAS blood commitment, we used single-cell RNA sequencing (scRNA-seq) of frozen PBMCs from 2 individuals in each patient group, using the 10X Chromium droplet-based platform. Unsupervised clustering based on gene expression identified 24 cell clusters among PBMCs, depicting the classical cell types found in the blood (**Extended Data Fig. 5a and 5b**). A cluster bias analysis performed by groups of patients strikingly showed a drastic loss of clusters derived from monocytes in VEXAS patients as compared to VEXAS-like, MDS and healthy controls (**Extended Data Fig. 5c and 5d**). Among all clusters, CD14^+^ CCL2^-^ and nonclassical CD14^lo^ CD16^+^ monocyte subsets were dramatically reduced in VEXAS patients (**Fig. 6a and 6b**). These results confirmed major differences observed using CyTOF and showing major reduction of nonclassical CD14^lo^ CD16^+^ monocyte fractions in VEXAS patients.

**Fig. 6.**
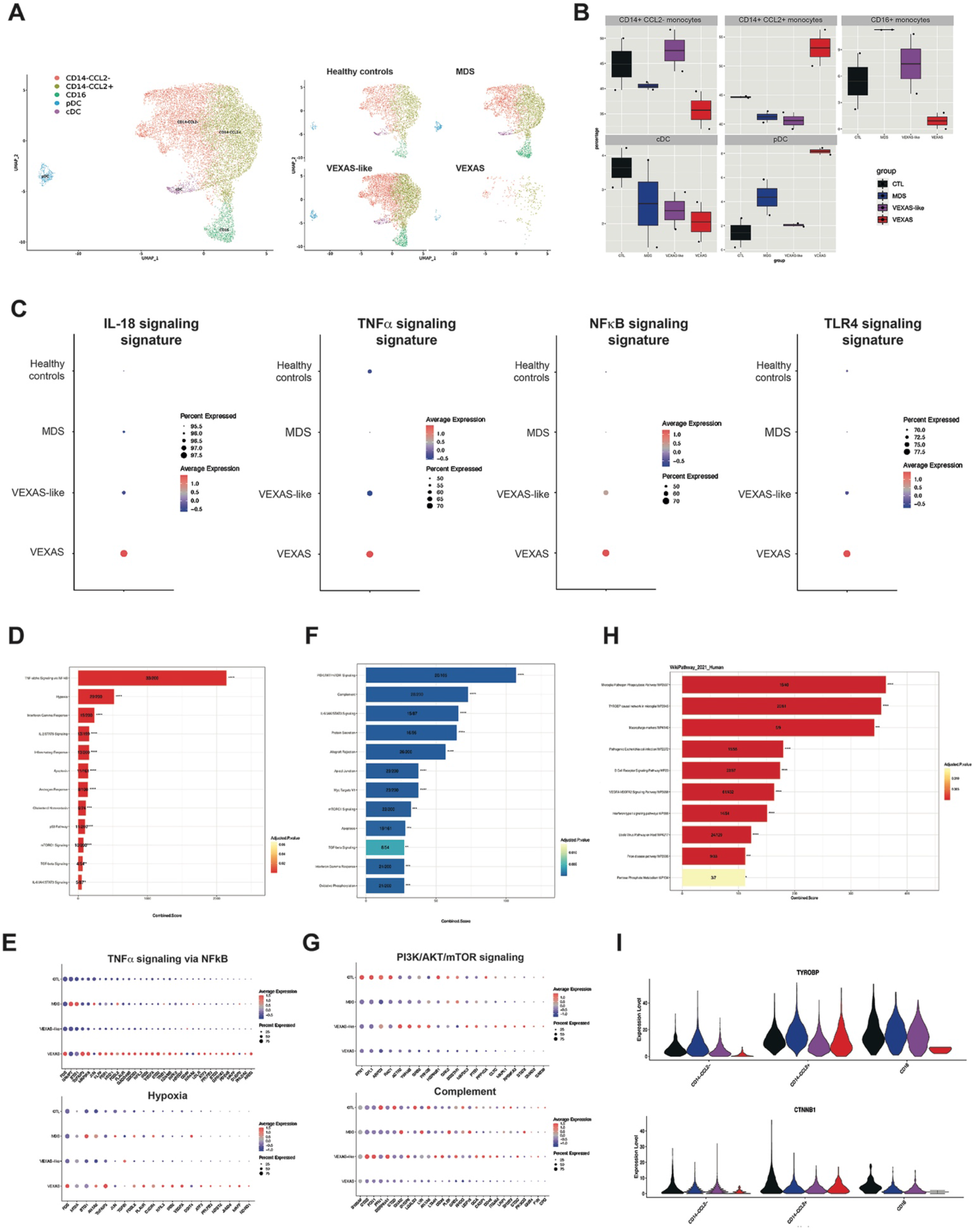
Visual representation of single-cell transcriptomic data of monocytes in VEXAS syndrome. (**A**) UMAP plots showing the projection of single myeloid cells from PBMCs from patients with VEXAS, VEXAS-like, MDS and healthy controls. (**B**) Proportion (frequencies) of the myeloid cell subsets, i.e. CD14+ CCL2+ monocytes, CD14+ CCL2-monocytes, CD16+ monocytes, cDC and pDC. (**C**) IL-18, TNF-α, NFκB and TLR4 signaling Gene expression signatures in monocytes from each patients’ group. The size of the dot represents the percentage of cells in the clusters expressing the gene expression signature and the color intensity represents the average expression of the signature in that cluster. (**D**) Enriched GO functions of up-regulated pathways in monocytes from VEXAS versus healthy controls. (**E**) Detailed analysis of the two most up-regulated pathways in monocytes from VEXAS, i.e. TNF-α signaling via NFκB pathway and hypoxia. (**F**) Enriched GO functions of up-regulated and down-regulated pathways in monocytes from VEXAS versus healthy controls. (**G**) Detailed analysis of the two most down-regulated pathways in monocytes from VEXAS, i.e. PI3K/AKT/mTOR signaling and complement pathways. (**H**) Top pathways enriched for pathway analysis through Wikipathways in dysregulated genes in monocytes from VEXAS versus healthy controls. (**I**) Expression levels in each monocyte subsets of *TYROBP*, encoding for DAP12, and *CTNNB1*, encoding catenin beta-1,in each patients’ group.

When testing molecular signatures from Nanostring Human Immunology at the single-cell level on PBMCs and monocytes, high expression of IL-18, TNF-α and NFκB signatures **(Extended Data Fig. 5e and Fig. 6c)** were observed as suggested by previous results **(Fig. 5c)**, as well as a TLR4 signaling signature, known to activate and signal downstream signaling pathways to activate NFκB and inflammasome complex. We next performed differential gene expression coupled with pathways analysis enrichment and identified specific up- and down-regulated pathways in monocytes from VEXAS patients. Among the most up-regulated pathways, we identified the TNF-α signaling via NFκB pathway with a very high expression of *FOS, TNFAIP3, MAP3K8* and *KLF9* in VEXAS samples, and hypoxia (**Fig. 6d and 6e**). Among the most down-regulated pathways, PI3K/AKT/mTOR signaling and complement pathways characterized circulating monocytes from VEXAS patients (**Fig. 6f and 6g**).

Moreover, molecular signatures obtained from Wikipathway highlighted new pathways potentially involved in the dramatic decrease of circulating monocytes, especially the *TYROBP* causal network in microglia (**Fig. 6h**). *TYROBP* and *CTNNB1* genes were drastically downregulated in the different monocytic clusters in VEXAS patients (**Fig. 6i**). Triggering Receptor Expressed on Myeloid cells 2 (TREM2), a cell surface receptor of the immunoglobulin superfamily that is expressed on microglia and myeloid cells, transmits intracellular signals through its adaptor, TYROBP/DAP12. A molecular defect of TYROBP/DAP12 in human monocytes was shown to deregulate the gene network pivotal for maintenance of myeloid cell function(Satoh et al., 2012). Also, TREM2/DAP12 signaling was shown to promote cell survival by activating the Wnt/β-catenin (*CTNNB1*) signaling pathway, at least in central nervous system (Zheng et al., 2017) and in bone homeostasis (Otero et al., 2012).

Finally, because of the upregulation of TNF-α family and NFκB signaling in whole blood and at the single-cell level, we investigated the contribution of proinflammatory programmed cell death in VEXAS. Plasma levels of lactate dehydrogenase (LDH), a marker of necrosis and cellular injury, correlated with VEXAS syndrome (**Fig. 1d**), as well as RIPK1 and RIPK-3, a key kinase involved in programmed necroptosis and inflammatory cell death (**Extended Data Fig. 6a**). Single-cell RNA sequencing of monocytes from VEXAS patients showed increased apoptosis, pyroptosis and necroptosis signatures in comparison with other groups, consistent with PANoptosis, an inflammatory programmed cell death pathway regulated by the PANoptosome complex with key features of pyroptosis, apoptosis, and/or necroptosis (Nguyen and Kanneganti, 2022) (**Extended Data Fig. 6b**). IRF1, a molecule long recognized for its roles in regulating cell death and as a key upstream regulator of PANoptosis, was significantly upregulated in peripheral blood of VEXAS patients (**Extended Data Fig. 6c**). Also, detailed signatures assessing the assembly of complexes involving RIPK1, i.e. complex 1, complex 2a and complex 2b, showed upregulation of all of these pathways (**Extended Data Fig. 6d)**. Importantly, upon activation of TNFR1, the complex 1, containing RIPK1, TNFR1-associated death domain (TRADD) and other signaling molecules, is rapidly formed and activates the induction of inflammatory and survival genes, and is subsequently followed by the assembly of complex 2a (ripoptosome) and 2b (necrosome) (Pasparakis and Vandenabeele, 2015; Wang et al., 2008).

Overall, these data confirm that monocytes from VEXAS patients display exaggerated proinflammatory signatures, UPR response and PANoptosis, contrasting with a defective TYROBP/DAP12 and β-catenin signaling pathway, modulating inflammation and cell death in the peripheral blood of VEXAS.

## DISCUSSION

By using high-throughput approaches on fresh blood samples, we were able to precise for the first time the inflammatory landscape of *UBA1*-mutated patients in comparison to adapted groups of controls, including inflammatory disorders without *UBA1* mutations. We have described unexplored areas of the myeloid/monocytic populations and have highlighted substantial differences in the abundance and phenotype of circulating monocytes in VEXAS patients. Peripheral blood monocytes from VEXAS patients showed dramatic decreased count and displayed many features of dysfunction and exhaustion, and aberrant expression of chemokine receptors. Correlation analysis of soluble mediators in peripheral blood also established that increased levels of markers of myeloid cells dysregulation and many proinflammatory cytokines as IL-1β and IL-18 were key features of VEXAS syndrome physiopathology. For the first time, these data suggest that the control of the undescribed inflammasome activation could be a specific feature and a potential therapeutic vulnerability of VEXAS.

According to our data obtained on blood and tissues compartment, we show that he dramatic decrease of monocytes we confirmed in VEXAS syndrome could be linked to an increased cell death and/or an enhanced migration into inflamed tissues.

In peripheral blood, bulk RNA sequencing focused on immune pathways confirmed many immune dysregulations and revealed an enrichment of TNF-α and NFκB signaling pathways. Moreover, Ingenuity Pathway Analysis identified the potential implication of FADD and RIPK1-dependent cell death and inflammation. Single-cell RNA sequencing analysis of peripheral monocytes confirmed the upregulation of these pathways at the single-cell level and their association to an increased UPR response signature and signatures related to cell death platform involving RIPK1. We also showed increased apoptosis, pyroptosis and necroptosis signatures, consistent with PANoptosis. PANoptosis has been increasingly implicated in infectious and inflammatory diseases, and the totality of biological effects in PANoptosis cannot be individually accounted for by pyroptosis, apoptosis, or necroptosis alone. The poor efficacy of conventional disease-modifying antirheumatic drugs and especially biologics targeting either IL-1β, TNF-α or IL-6 to control VEXAS could support the concomitant engagement of different modes of programmed cell death especially when used alone or sequentially (Gullett et al., 2022). These high-throughput analyses also highlight a defective TYROBP/DAP12 and β-catenin signaling pathway in pathological monocytes which could explain a lack in survival signals, especially in monocytes from VEXAS. Future works will be needed determine whether targeting cell death, especially RIPK1-dependent pathway and PANoptosome complex, can be used as a therapeutic strategy in VEXAS. A monitoring of these dysregulations would be investigated in vivo and in vitro to appreciate the efficacy of treatments.

A strength of this study is the possibility to get some information about the fate of myeloid cells in tissues that seems different to their behavior in peripheral blood. As expected, bone marrow analysis shows an overrepresentation of the myeloid cell lineage with mature and immature forms which contrasts with observations in peripheral blood. Dysregulation of the bone marrow microenvironment was showed in the pathophysiology of many myeloid disorders by influencing the behavior of the stem cell compartment (Yamashita et al., 2020). The role of the bone marrow microenvironment in VEXAS syndrome is yet totally unknown and its exploration should be an important field of investigation especially in terms of proinflammatory molecules secretion by stromal cells and survival factors for mutated cells. In the skin from VEXAS patients, we showed abundant monocytes and macrophages, with only very few infiltrating monocytes that colocalized with cleaved caspase-3, a marker of active cell death. High expression of chemokines receptors by circulating monocytes supported by CyTOF data and increased levels of chemokines, especially the CCL2/CCR2 axis, also support an enhanced migration of monocytes into tissues. Future studies will be necessary to identify the signals which allow the skin infiltration by mutated cells and to determine the potential protective role of this environment. If specific pathways are involved in this propagation of the mutated cells, some therapeutic options should be considered in addition to topic treatment.

Our study has however some limitations. The difficulty to isolate sufficient circulating monocytes from VEXAS patients did not allow us so far to evaluate more precisely the mechanisms driving their important decrease, especially cell death pathways. Future experiments on sorted monocytes from VEXAS and controls, on genetically modified monocytic cell lines or on bone marrow-derived monocytes will be needed to provide new data on the biology of mutated cells.

Although future works and clinical trials that can provide additional insights and an evaluation of targeting cell death and inflammasome pathways in VEXAS have yet to be completed, based on our observations, such pathways could be highlighted as therapeutic vulnerabilities. The role of a residual normal hematopoiesis in VEXAS patients and the evolution of the circulating monocytes under treatments will also represent major points to monitor in prospective clinical trials. Moreover, in a near future, these approaches will be used to usefully study VEXAS-like disorders whom genetic background is not yet elucidated.

## ONLINE CONTENT

Any methods, additional references, reporting summaries, source data, extended data, supplementary information, acknowledgements, peer review information; details of author contributions and competing interests; and statements of data and code availability are available online.

## METHODS

No statistical methods were used to predetermine sample size. The experiments were not randomized, and the investigators were not blinded to allocation during experiments and outcome assessment. Our research complies with all relevant ethical regulation, as detailed in the ‘Cohorts’ section.

### Cohort

This non-interventional study was conducted between January, 2021 and May, 2021, in the setting of the local RADIPEM biological samples collection derived from samples collected in routine care. Biological collection and informed consent were approved by the Direction de la Recherche Clinique et Innovation (DRCI) and the French Ministry of Research (N°2019-3677). A written informed consent was collected for all participants. None of the study participants received compensation. This study was conducted in compliance with the Good Clinical Practice protocol and the principles of the Declaration of Helsinki, and received approval from the Cochin Hospital Institutional Review Board (number AAA-2021-08040).

Patients included in this study were: adults patients aged over 18 years old, with a history of inflammatory syndrome characterized by relapsing polychondritis, Sweet’s syndrome, polyarteritis nodosa, or other unclassified inflammatory disease, and associated or not with well-defined hematologic condition (mainly myelodysplastic syndrome). Epidemiological, demographic, clinical, laboratory, treatment, and outcome data were extracted from medical charts using a standardized data collection form. At time of sampling and according to the pandemic situation, all included patients were tested for SARS-CoV-2 infection and were all negative.

For VEXAS, VEXAS-like and MDS patients, genomic DNA was extracted from blood samples and the third exon of the gene was analyzed by Sanger approach to detect the described mutations using specific primers as previously reported (Forward 5’-TCCAAAGCCGGGTTCTAACT-3’ / Reverse 5’-GGGTGTGCAGTAGGGAAAAA-3’) ^2^.

### Routine plasmatic biomarkers measurements

Routine blood examinations were complete blood count and plasmatic biochemical tests, including C-reactive protein (CRP), ferritin, interleukin-(IL)-6, calprotectin and lactate dehydrogenase (LDH). Plasma EDTA were collected and frozen at -80°C for subsequent measurement of protein biomarkers. Lactate-deshydrogenase (LDH) concentrations were measured using the LDHI2 enzymatic UV assay on a cobas c701 analyzer (Roche Diagnostics Meylan, France). The measuring range of the assay extended from 10 to 1,000 UI/L. CRP concentrations were measured using the Tina-quant CRP-Gen3 immunoturbidimetric assay on a cobas c701 analyzer (Roche Diagnostics Meylan, France). The measuring range of the assay extended from 0.5 to 15 mg/L. Physicians in charge of the patients were blinded to the results of biomarkers, and biologists were blinded to the emergency diagnosis suspected by physicians.

### Multiparameter phenotyping of peripheral blood leukocytes using mass cytometry

The Maxpar^®^ Direct™ Immune Profiling System (Fluidigm, Inc Canada) was used for high-dimensional immune profiling of whole blood, using a 30-marker antibody panel with the addition of eleven markers: anti-CD11b conjugated to 106Cd, anti-CD64 conjugated to 111Cd, anti-CCR3 conjugated to 113Cd, anti-FcERI conjugated to 116Cd, anti-CD69 conjugated to 142Nd, anti-CD117 conjugated to 165Ho, anti-CD336/NKp44 conjugated to 169Tm, anti-Tim3 conjugated to 159Tb (1 μg/μL concentration), anti-CD335/NKp46 conjugated to 162Dy (1 μg/μL concentration), anti-PD-1 conjugated to 175Lu (1 μg/μL concentration) and anti-PD-L1 conjugated to 209Bi (1 μg/μL concentration). For each sample, 10 μL of 10 KU/mL heparin solution was added to 1 mL of whole blood, then the samples were incubated for 20 minutes at room temperature. After incubation, 270 μL of heparin treated whole blood was directly added to the dry antibody cocktail and additional antibodies were added, for an incubation of 30 minutes. Red blood cells were then lysed immediately using CAL-Lyse Lysing Solution (Life Technologies). After a 10-min incubation, 3 mL of MaxPar Water was added to each tube for an additional 10-min incubation. Cells were washed three times using MaxPar Cell Staining Buffer and then fixed for 10 min using with 1.6% paraformaldehyde (Sigma-Aldrich, Lyon, France). Cells were washed once with MaxPar Cell Staining Buffer and incubated one hour in Fix and Perm Buffer with 1:1000 of Iridium intercalator (pentamethylcyclopentadienyl-Ir (III)-dipyridophenazine, Fluidigm, Inc Canada). Cells were washed and resuspended in Maxpar Cell Acquisition Solution, a high-ionic-strength solution, at a concentration of 1 million cells per mL and mixed with 10% of EQ Beads immediately before acquisition. Cell events were acquired on the Helios mass cytometer and CyTOF software version 6.7.1014 (Fluidigm, Inc Canada) at the “Plateforme de Cytométrie de la Pitié-Salpetriere (CyPS).” An average of 400,000 events were acquired per sample. Mass cytometry standard files produced by the HELIOS were normalized using the CyTOF Software v. 6.7.1014. This method normalizes the data to a global standard determined for each log of EQ beads.

FCS3.0 files generated by the Helios were analyzed using GemStone software (Verity Software House, Topsham, ME), an automated analysis system. This system is integrated with dimensionality-reduction mapping known as Cauchy Enhanced Nearest-neighbor Stochastic Embedding (Cen-se™), which generates a visual display of high-dimensional data labeled with the major cell populations.

The multiparametric analysis of activation and immune checkpoint markers was performed on FlowJo and the data generated were then analysed using Tableau Desktop. For viSNE analysis (Cytobank Inc, Mountain View, CA, USA), mapping integrating a total of 50,000 PBMCs sampled and analyzed for each group FCS file. Parameter viSNE maps were created with all markers. The settings used for the viSNE run were as follow: iterations (3000), perplexity (70) and theta (0.5). ViSNE maps are presented as means of all samples in each disease severity category.

### Imaging mass cytometry

Tissue sections were dewaxed in xylene for 20 min. Samples were rehydrated in a graded series of alcohol (ethanol:deionized water 100:0, 95:5 and 70:30; 5 min each). After that, slides were washed in Maxpar Water^®^ for 5 minutes in a Coplin jar placed on an orbital shaker plate with gentle agitation. Slides were put into to Tris-EDTA buffer at pH 8 followed by heat-induced epitope retrieval in a pressure cooker (Medite) for 25 min at 92 °C. Samples were left to cool in the Tris-EDTA buffer for 20 min followed by cooling in Tris-buffered saline (TBS) at room temperature for at least 20 min.

After that, slides were washed in Maxpar Water^®^ for 10 minutes in a Coplin jar placed on an orbital shaker plate with gentle agitation, then in Maxpar PBS^®^ for 10 minutes. Slides were then blocked with blocking buffer containing 3% BSA in Maxpar PBS^®^ for 45 min. Samples were then stained with antibody mix (diluted in blocking buffer) and incubated overnight at 4 °C. Samples were washed three times in 0.2% Triton X-100 in Maxpar PBS^®^ for 8 minutes with slow agitation in Coplin jars, then in Maxpar PBS^®^ for 5–10 min, before incubating for 30 min with iridium intercalator diluted in Maxpar PBS^®^ followed by three washes in Maxpar PBS^®^ (5 min each wash). Samples were then air dried before IMC acquisition.

Data acquisition was performed on a Helios time-of-flight mass cytometer (CyTOF) coupled to a Hyperion Imaging System (Fluidigm). Selected areas for ablation were larger than the actual area of interest to account for loss of overlapping areas among sections due to cumulative rotation. The selected area ablated per section was around 1 mm^2^. To ensure performance stability, the machine was calibrated daily with a tuning slide spiked with five metal elements (Fluidigm). All data were collected using the commercial Fluidigm CyTOF software v.01. Finally, regions of interest with markers of interest (DNA1, smooth muscle actin [SMA], CD3, CD4, CD8, CD14, CD16, CD31, CD68 and CD163) were visualized using MCD™ Viewer v1.0.560.6 software.

For each sample, genomic DNA was extracted and the mutational status of UBA1 was tested as previously reported. The VEXAS skin biopsies presented the same UBA1 mutation that was identified in fresh blood samples.

### Cytokine assays

Plasma were analyzed using the Luminex xMAP technology and a 45 plex assay (R&D Systems). To quantify IFN-α, IFN-γ and IL-17A at ultrasensitive concentrations, Single Molecule array (Simoa) technology was used and a homebrew triplex assay was developed as previously described (Bondet et al, 2021). All the assays were run on a 2-step configuration on the Simoa HD-1 Analyzer (Quanterix, US). Limit of detection (LoD) was defined (using the highest bottom value of 95% CI in Prism 9). The R package “limma” v3.44.3 was used to correct for Luminex kits batch effect. PCA and violin plots were performed using the R packages “factoextra” v1.0.7 and “ggplot2” v3.3.3 respectively. Interleukin-6 concentrations were measured using the IL-6 ECLIA assay on a cobas E801 analyzer (Roche Diagnostics Meylan, France). The measuring range of the assay extended from 0.5 to 5,000 ng/L. Calprotectin concentrations were measured using the turbidimetric assay (Gentiane) on a cobas c501 analyzer (Roche Diagnostics Meylan, France). The measuring range of the assay extended from 0.5 to 15 mg/L. All quality controls during the study were conformed to. IL-1β (Human IL-1 beta/IL-1F2 Quantikine ELISA Kit, Catalog # DLB50, R&D Systems), IL-18 (Human Total IL-18/IL-1F4 Quantikine ELISA Kit, Catalog # DL180, R&D Systems) and galectin-3 (Human Galectine-3 Quantikine ELISA Kit, Catalog # DGAL30, R&D Systems) plasma levels were measured using an ELISA kit and following the manufacturer’s instructions.

### Gene expression analysis

Total RNA was extracted from 2 ml of whole blood collected on EDTA using the Promega Maxwell LEV Simply RNA Kit (Ref AS1280). RNA concentrations were measured using a Nanodrop One (Thermo Scientifics). Total RNA samples were analyzed using the Human Immunology kit v2 panel profiling 594 immunology-related human genes according to manufacturer’s instructions (Nanostring). Gene expression data were normalized using control probes and housekeeping genes selected using the geNorm method of the Advanced Analysis package (nSolver Analysis Software 4.0). As gene expression was measured using two different batches of CodeSets, batch effect correction was performed using the removeBatchEffect function of the R package “limma” v3.44.3.

To identify gene expression differences between groups, multiple linear regression was performed while correcting for lymphocyte, neutrophil and monocyte cell proportions to control for potential gene expression differences due to cellular differences between patients. The analysis was implemented using the R package “broom”‘ v0.7.5. Multiple testing correction was then applied to select the significant genes. PCA, volcano plots and heatmaps were performed using the “factoextra” v1.0.7, “EnhancedVolcano” v1.6.0 and “pheatmap” v1.0.12 respectively. Finally, gene signatures (Table x) were calculated for each sample by using the zScore function of the R package multiGSEA v1.1.99 and gene set enrichment analysis was performed as previously described (Hadjadj et al, Science, 2020).

### Single-cell RNA sequencing

The scRNA-seq libraries were generated using Chromium Single Cell Next GEM 3′ Library & Gel Bead Kit v.3.1 (10x Genomics) according to the manufacturer’s protocol. Briefly, cells were counted, diluted at 1000 cells/µL in PBS+0,04% and 20 000 cells were loaded in the 10x Chromium Controller to generate single-cell gel-beads in emulsion. After reverse transcription, gel-beads in emulsion were disrupted. Barcoded complementary DNA was isolated and amplified by PCR. Following fragmentation, end repair and A-tailing, sample indexes were added during index PCR. The purified libraries were sequenced on a Novaseq (Illumina) with 28 cycles of read 1, 8 cycles of i7 index and 91 cycles of read 2.

Sequencing reads were demultiplexed and aligned to the human reference transcriptome (GRCh38-2020-A directly download from 10x), using the CellRanger Pipeline (v-5.0.1). The unfiltered raw UMI counts from cellranger were loaded inot Seurat v4.0.3 (Stuart et al., 2019) for quality control, data integration and downstream analyses. Doublets, empty sequencing beads and apoptotic cells were removed by filtering out cells with fewer than 500 features or a mitochondrial content higher than 20%. Data from each sample were normalized and scaled using the sctransform method, and batch effect between samples was corrected using Seurat’s FindIntegratedAnchors.

On this integrated dataset, we computed the principal component analysis on the 2000 most variable genes. UMAP was carried out using the 20 most significant PCs, and community detection was performed using the graph-based modularity-optimization Louvain algorithm from Seurat’s FindClusters function with a 0.8 resolution.

Cell type labels were assigned to resulting clusters based on a manually curated list of marker genes as well as previously defined signatures of the well-known PBMC subtypes (Monaco et al., 2019). All clusters were annotated, and 63630 cells were kept for further analysis.

Differential expression was performed on different groups, using the FindMarkers function of Seurat on the RNA assay with default parameters (Wilcoxon testing with Bonferroni correction). Only genes with adjusted p-values < 0.05 were selected as significant. The lists of differentially expressed genes were further divided into UP and DOWN regulated genes based on the avg_log2FC; avg_log2FC >0 for the UP regulated genes and and avg_log2FC <0 for the DOWN regulated ones.

The signature scores were calculated using the function AddModuleScore from Seurat, and dot plots were used to visualise the change in signature signal between conditions. The data for the analysis on the myeloid and monocyte populations were obtained using the subset function from Seurat.

### Statistical analysis

CyTOF data were analyzed with FlowJo version 10 software. Calculations were performed using Excel 365 (Microsoft). Figures were drawn on Prism 9 (GraphPad Software). Statistical analysis was conducted using GraphPad Prism 9. P values were determined by a Kruskal-Wallis test, followed by Dunn’s post-test for multiple group comparisons with median reported; *P < 0.05; **P < 0.01; ***P < 0.001. All tests were two-sided. Correlation matrices of cytokines from patient blood were calculated. Only significant correlations (<0.05) are represented in color.

## Data Availability

All data supporting the findings of this study are available within the article or from the corresponding authors upon reasonable request without any restrictions.

## ACKNOWLEGMENTS

We acknowledge all people that support the Fonds IMMUNOV, for Innovation in Immunopathology. We also acknowledge health care workers involved in the diagnosis and treatment of patients. We thank all the patients, supporters, and our families for their confidence in our work.

We also thank the following colleagues for their collaboration for the diagnosis of VEXAS syndrome: Jean-Sébastien Allain, Adrien Bigot, Boris Bienvenu, Didier Bouscary, Weniko Care, Benjamin de Sainte-Marie, Justine Decroocq, Benoit Faucher, Edouard Flamarion, Cécile Golden, Sylvie Grosleron, Helder Gil, Julie Graveleau, Sébastien Humbert, Nathalie Jacque, Marie Kostine, Valentin Lacombe, Dan Lipsker, Nadine Magy-Bertrand, Aurore Meyer, Marielle Roux-Sauvat, Maxime Samson, Julie Seguier, Laure Swiader, Sylvain Thépot, Olivier Tournilhac, Marguerite Vignon, Julien Vinit, Stéphane Vinzio.

We also acknowledge Morgane Le Gall for her help for the Ingenuity Pathway Analysis.

## AUTHORS CONTRIBUTIONS

Experimental strategy and design: O.K., C.P., M.M., D.D. and B.T. Laboratory experiments: O.K., C.P., M.T., A.C., M.L., E.D., B.B., P.S., H-K.E., M.M., D.D. and B.T. Cohort management and clinical research: O.K., C.P., M.T., E.D., M.M., D.D. and B.T. Statistical analyses: O.K., C.P., M.T., A.C., F.C., J.B., M.M., D.D. and B.T. Manuscript writing: O.K. and B.T. Manuscript editing: O.K., C.P., M.T., A.CM.M., D.D. and B.T.

All authors critically revised the manuscript for important intellectual content and gave final approval for the version to be published. All authors agree to be accountable for all aspects of the work in ensuring that questions related to the accuracy or integrity of any part of the work are appropriately investigated and resolved.

## ETENDED DATA

**Extended data Fig. 1.**
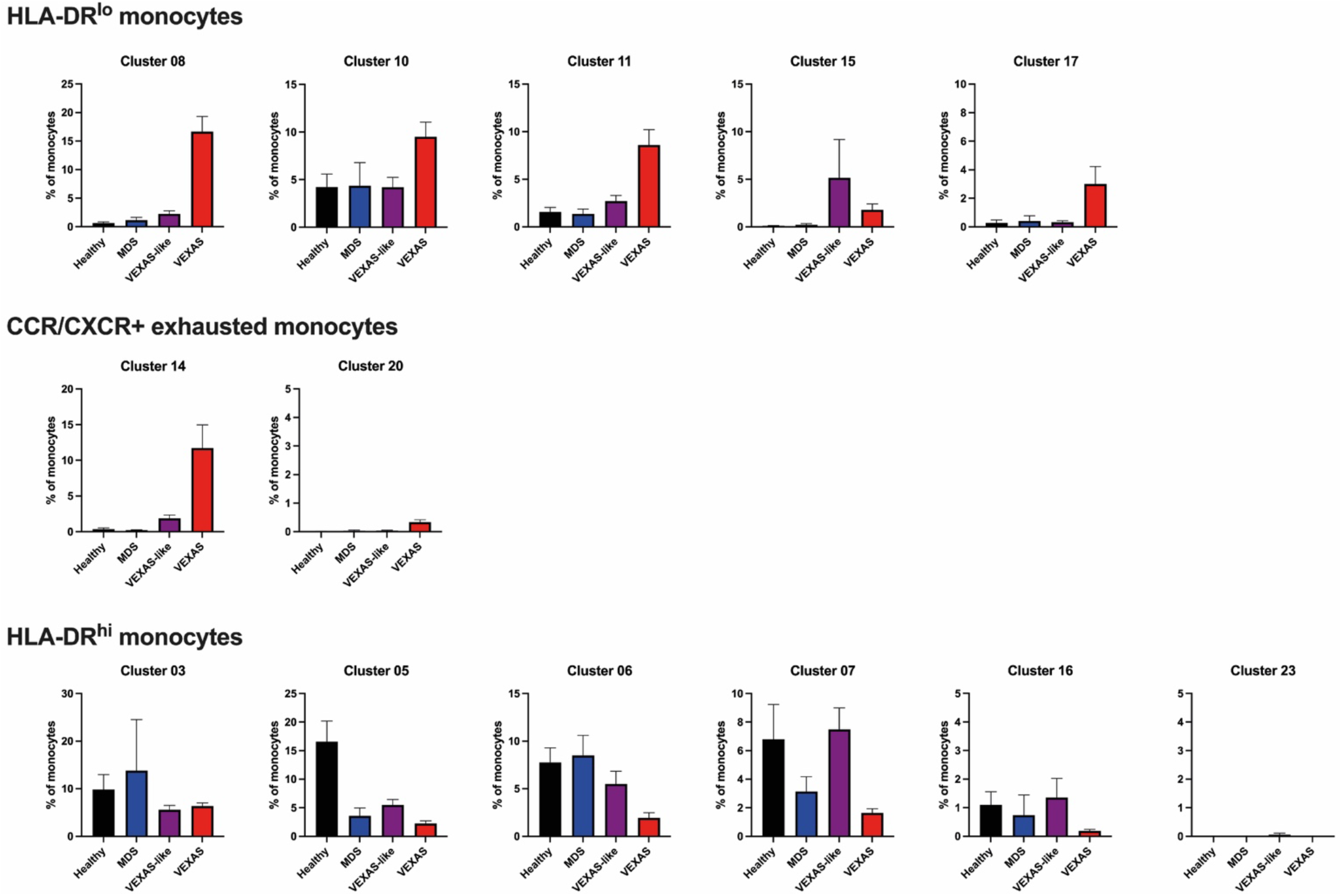
Distribution of monocyte clusters among whole monocytes in patients with VEXAS syndrome, VEXAS-like, MDS and healthy controls. Clusters 8, 10, 11, 15 and 17, corresponding to HLA-DR^lo^ monocytes (detailed in upper panel), and clusters 14 and 20 (middle panel), corresponding to exhausted monocytes expressing chemokine receptors, are increased in VEXAS. Clusters 3, 5, 6, 7 and 16 (lower panel), corresponding to HLA-DR^hi^ monocytes, are decreased in VEXAS.

**Extended data Fig. 2.**
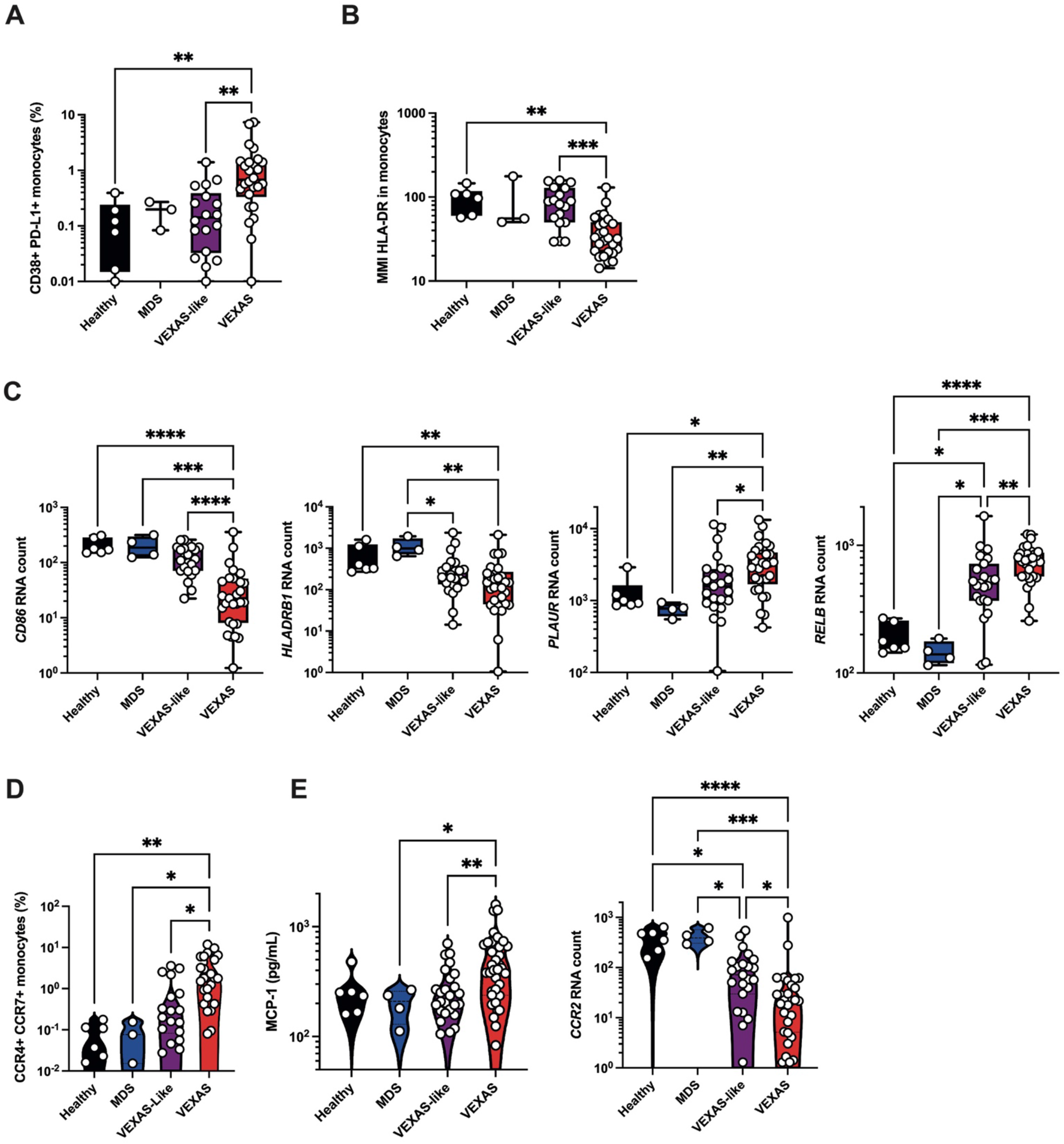
Activation status of monocytes at cellular and molecular level. Each dot represents a single patient studied as described by CyTOF, cytokines measurement or RNA studied by Nanostring nCounter analysis. **(A)** Analysis of exhausted monocytes based on the expression of CD38 and PD-L1 by mass cytometry. Each dot represents a single patient. **(B)** Cell surface expression of HLA-DR on monocytes, evaluated by mean metal intensity (MMI). **(C)** Absolute RNA counts using Nanostring nCounter technology and showing decreased expression of CD86 and HLADRB1, and increased expression of PLAUR and RELB transcripts in VEXAS in comparison to other groups. **(D)** Proportion (frequencies) of CCR4+ CCR7+ expressing monocytes. **(E)** CCL2/MCP-1 protein plasma concentration measured with Luminex technology, and absolute RNA count for CCR2. Each dot represents a single patient. RNA data are extracted from the Nanostring nCounter analysis. P values were determined by the Kruskal-Wallis test, followed by Dunn’s post test for multiple group comparisons. *P <0.05; **P <0.01; ***P <0.001, ****P <0.0001.

**Extended data Fig. 3.**
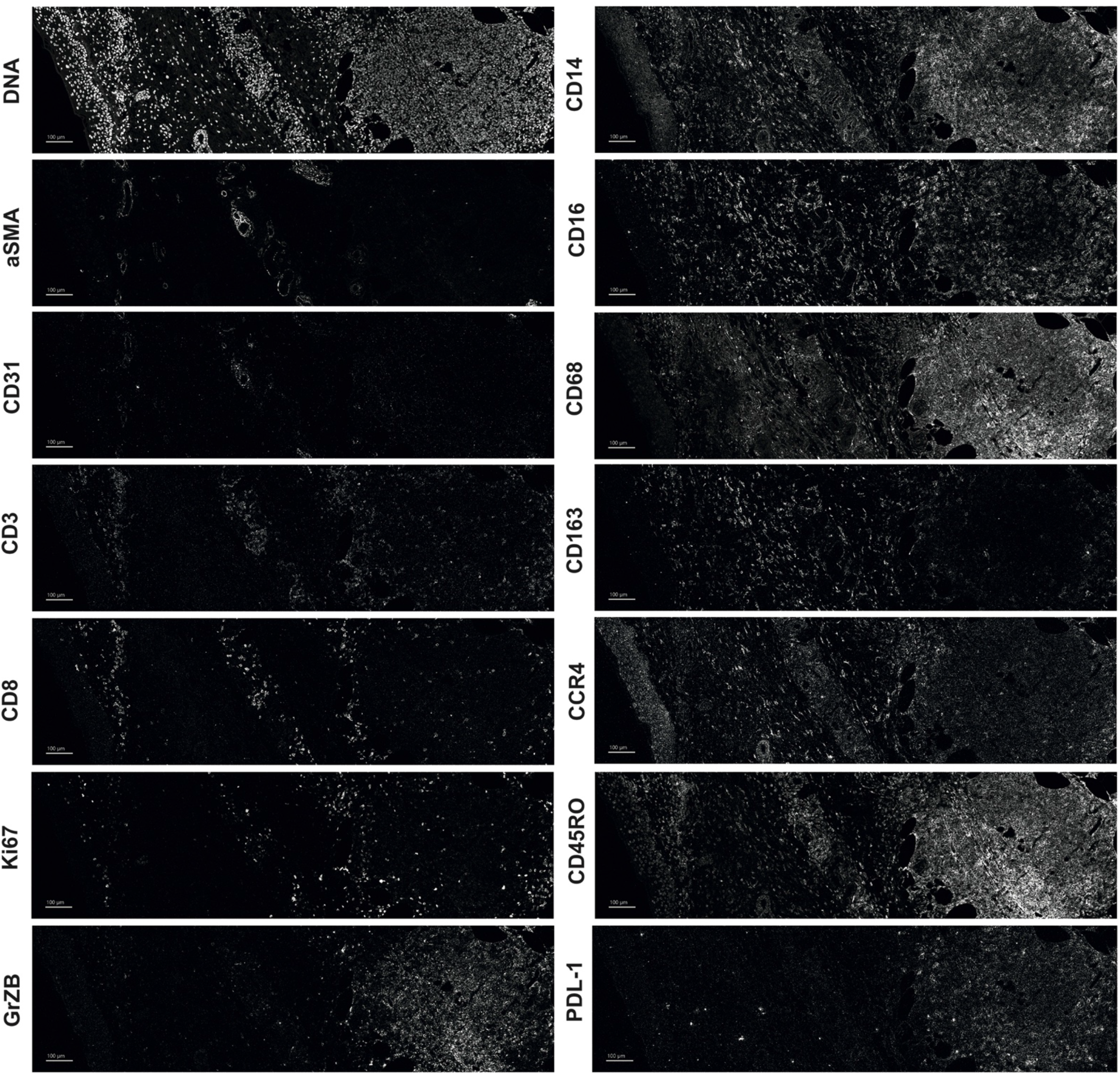
Expression of cell markers on pathological skin lesion from a VEXAS patient using imaging mass cytometry. Imaging mass cytometry on skin biopsy showing expression of DNA, alpha-smooth muscle actin (aSMA), CD31, CD3, CD8, CD14, CD16, CD68, CD163, CCR4, Ki67, CD45RO, granzyme B (GrZB) and PDL-1. Illustrative pictures are shown.

**Extended data Fig. 4.**
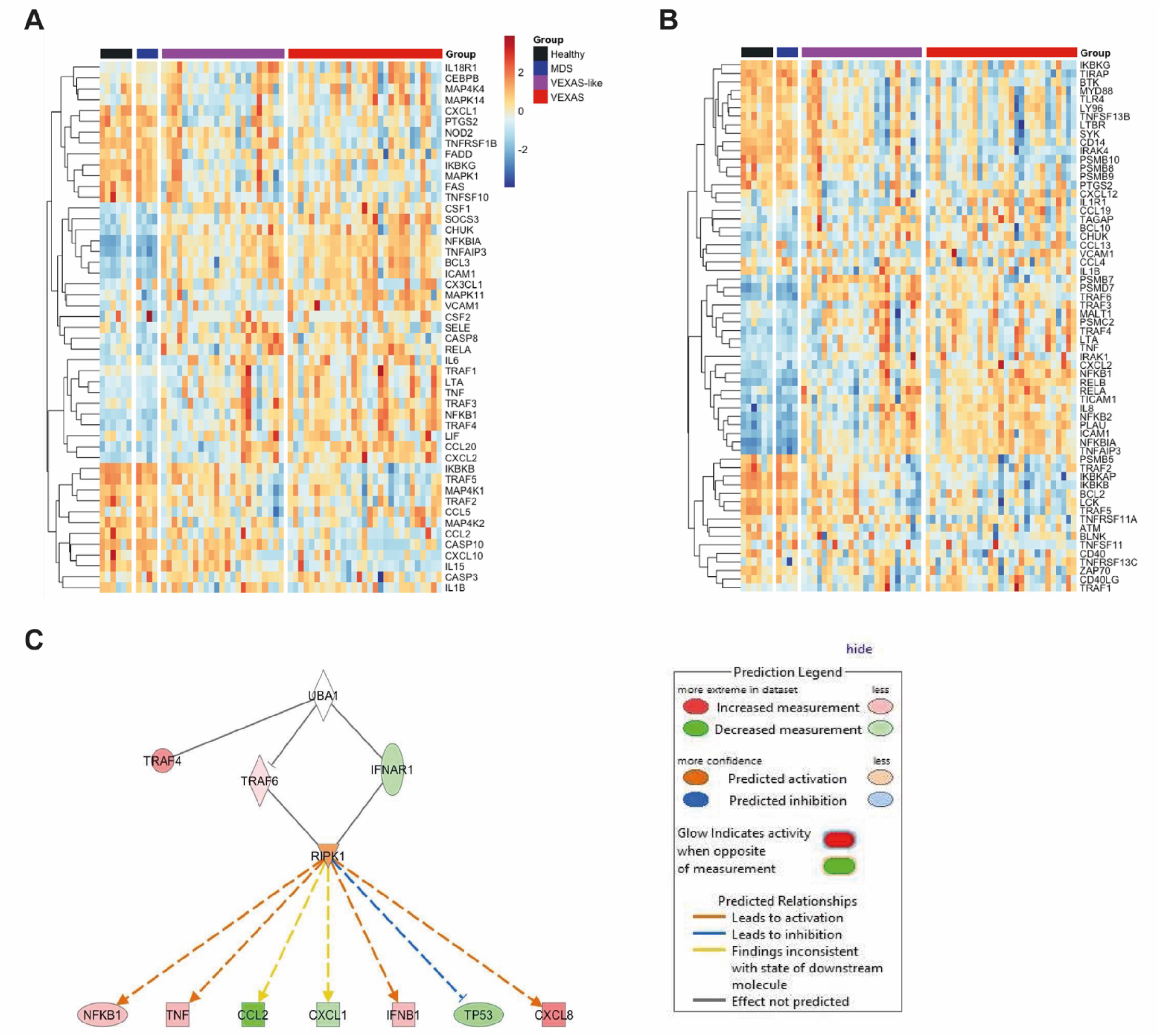
TNF-α pathway signaling and NFκB signaling signatures in patients with VEXAS syndrome, VEXAS-like, MDS and healthy controls. **(A)** Heatmap representation of genes encoding proteins involved in TNF-α pathway signaling in each group. **(B)** Heatmap representation of genes encoding proteins involved in NFκB signaling in each group. **(C)** Ingenuity Pathway Analysis studying connections between differentially regulated genes, upstream regulators and signaling pathways potentially responsible for the observed differences in gene expression, showing the role of RIPK1 involved into the ripoptosome pathway in VEXAS samples comparing to healthy controls.

**Extended data Fig. 5.**
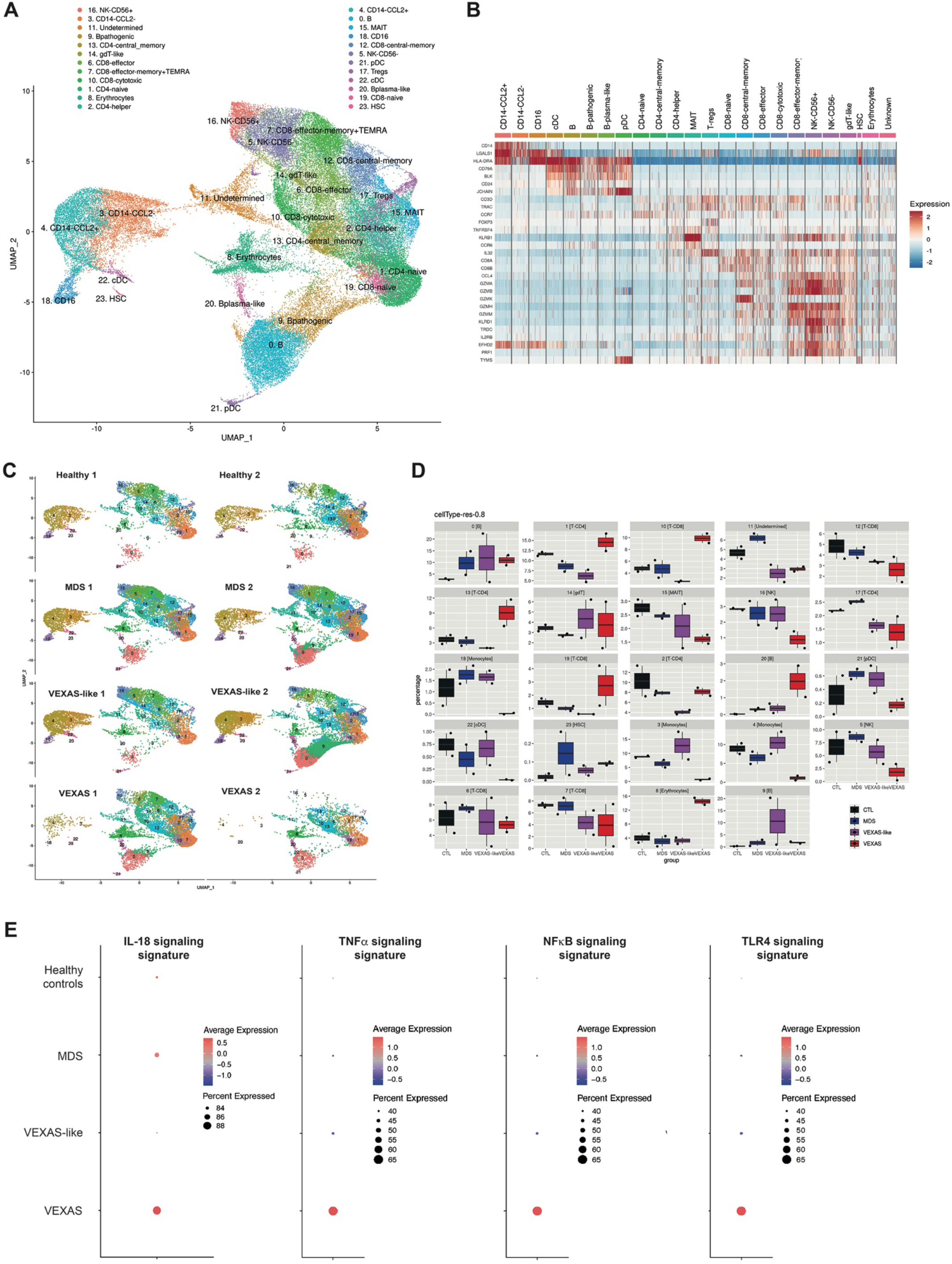
Visual representation of single-cell transcriptomic data of PBMCs in VEXAS syndrome. (**A**) UMAP plots showing the projection of cell populations identified from PBMCs. (**B**) Heatmap of the normalized top differently expression genes in the different cell populations in PBMCs. (**C**) UMAP plots showing the projection of all PBMCs from patients with VEXAS, VEXAS-like, MDS and healthy controls. (**D**) Proportion (frequencies) of all different cell populations identified in the PBMCs of patients. (**E**) IL-18, TNF-α, NFκB and TLR4 signaling gene expression signatures in PBMCs from each patients’ group. The size of the dot represents the percentage of cells in the clusters expressing the gene expression signature and the color intensity represents the average expression of the signature in that cluster.

**Extended data Fig. 6.**
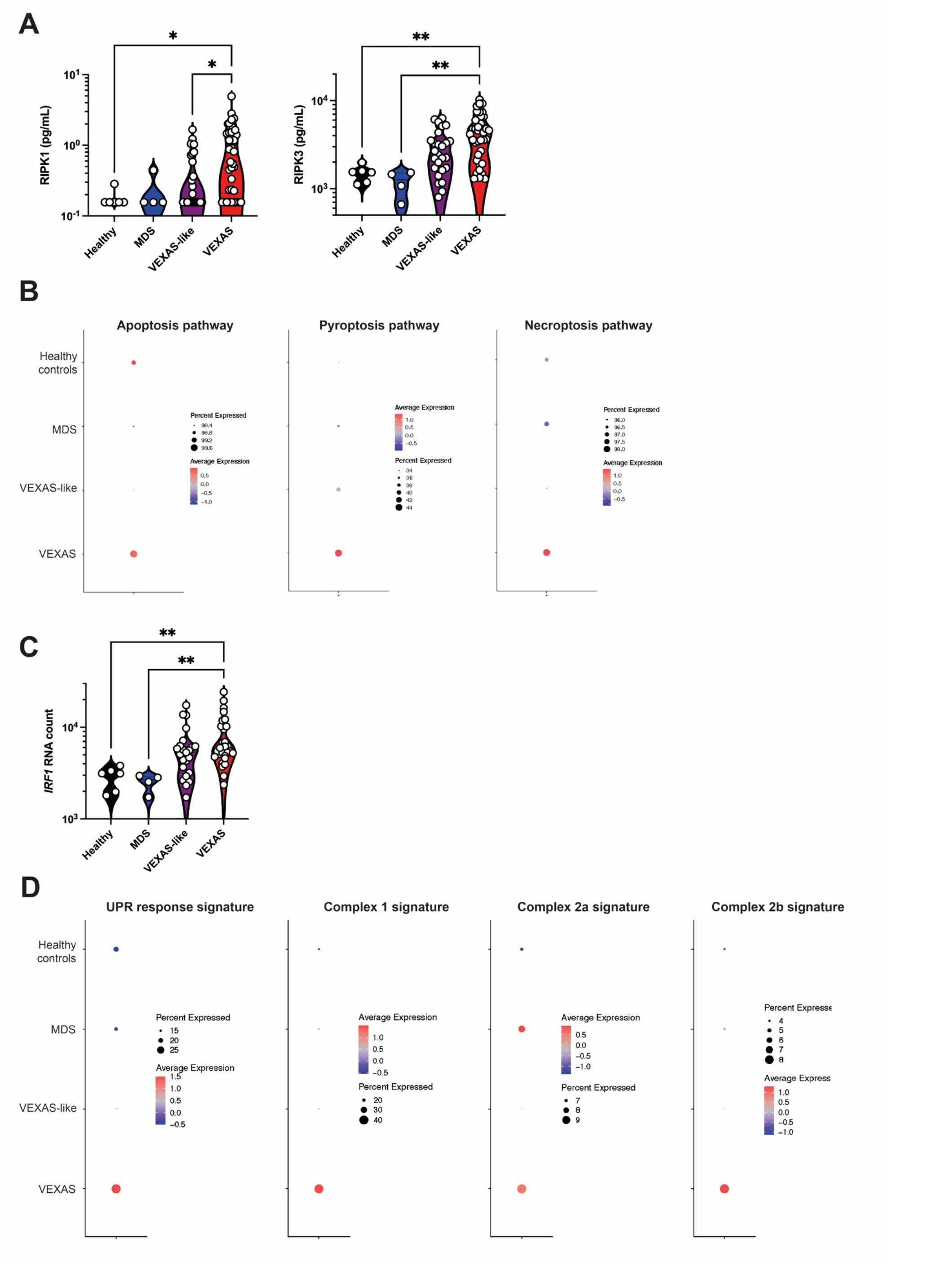
Inflammatory programmed cell death pathway in monocytes from VEXAS syndrome. (**A**) RIPK1 and RIPK3 plasma concentrations in patients with VEXAS, VEXAS-like, MDS and healthy controls. Each dot represents a single patient. (**B**) Apoptosis, pyroptosis and necroptosis gene expression signatures in monocytes from each patients’ group. The size of the dot represents the percentage of cells in the clusters expressing the gene expression signature and the color intensity represents the average expression of the signature in that cluster. **(C)** Absolute RNA counts using Nanostring nCounter technology and showing decreased increased of *IRF1* transcripts in VEXAS. Each dot represents a single patient. (**D**) Signatures assessing the URP response and assembly of complexes involving RIPK1, i.e. complex 1, complex 2a and complex 2b, in monocytes from each patients’ group. P values were determined by the Kruskal-Wallis test, followed by Dunn’s post test for multiple group comparisons. *P <0.05; **P <0.01.

## Supplementary Tables

**Supplementary Table 1.**
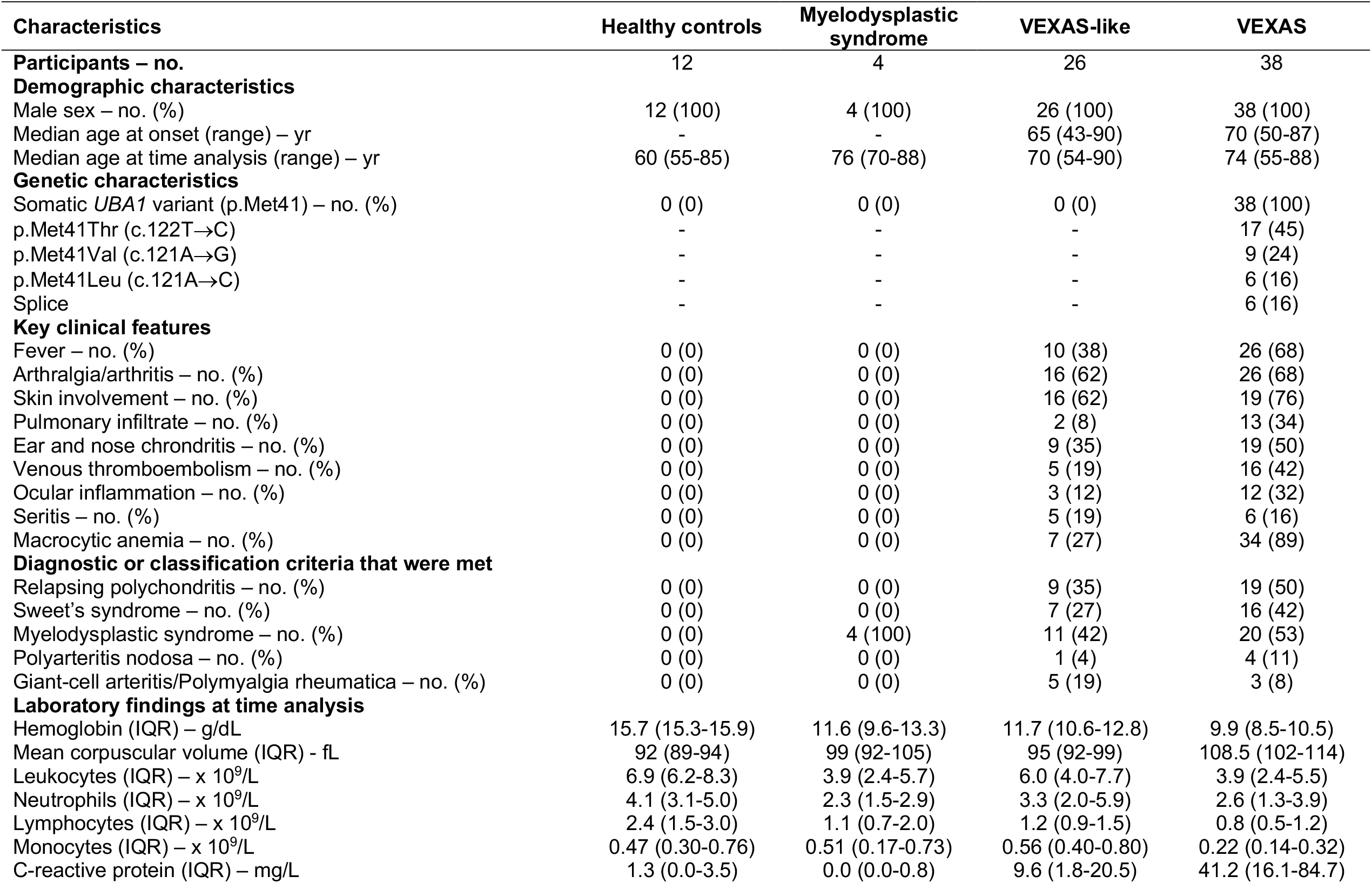
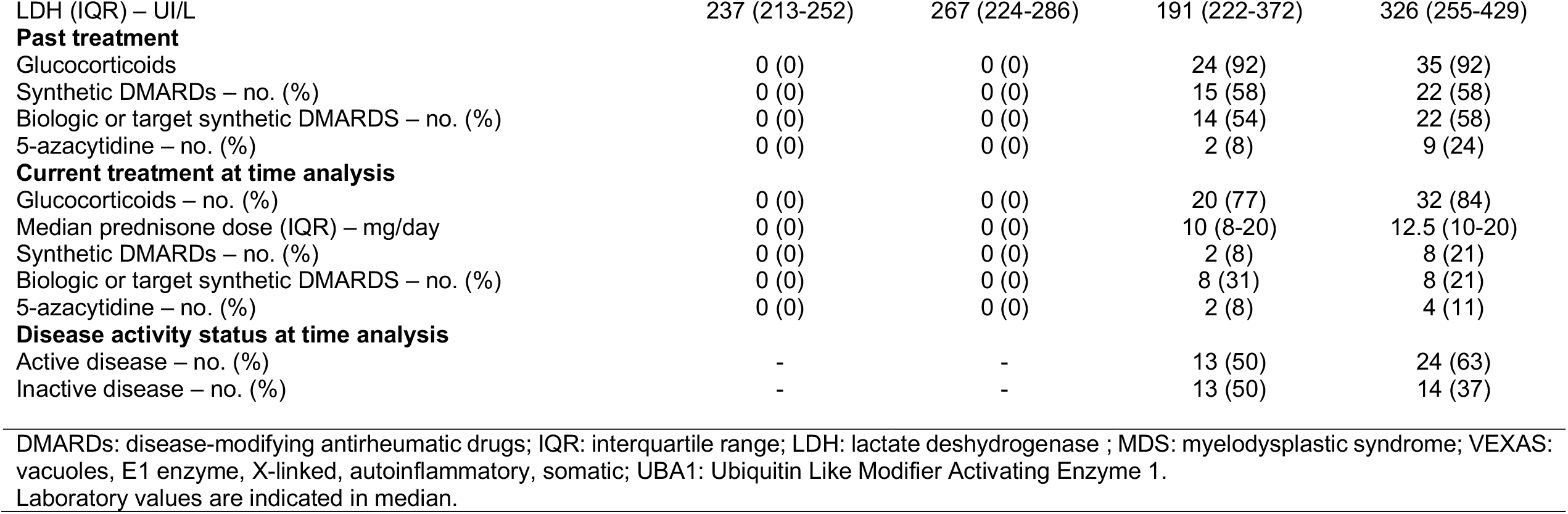
Demographic and clinical characteristics of participants included in the study.

**Supplementary Table 2.** Phenotypic definition of immune cell populations.

|  |  |
| --- | --- |
| **CD8+ T cells** | CD3+ CD66b- CD19- CD8+ CD4- CD14- CD161- TCR$\gamma\delta$- CD123- CD11c- |
| **CD8 differentiation stages** |  |
| *Naive* | CCR7+ CD27+ CD45RA+ |
| *Central memory* | CCR7+ CD27+ CD45RA- |
| *Effector memory* | CCR7- CD27+ |
| *Terminal effector* | CCR7- CD27- |
| **CD4+ T cells** | CD3+ CD66b- CD19- CD8- CD4+ CD14- CD161- TCR$\gamma\delta$- CD123- CD11c |
| **CD4 differentiation stages** |  |
| *Naive* | CCR7+ CD27+ CD45RA+ |
| *Central memory* | CCR7+ CD27+ CD45RA- |
| *Effector memory* | CCR7- CD27+ CD45RA- |
| *Terminal effector* | CCR7- CD27- CD45RA- |
| **CD4 polarization stages** |  |
| *Th1* | CXCR3+ CCR6- CCR4- CXCR5- |
| *Th2* | CXCR3- CCR6- CCR4+ CXCR5- |
| *Th17* | CXCR3- CCR6+ CCR4+ CXCR5- |
| *Regulatory T cells* | CD25+ CD127- CCR4+ HLA-DR- |
| **$\gamma\delta$ T cells** | CD3+ CD66b- CD14- CD8dim CD4- TCR$\gamma\delta$+ |
| **Natural killer (NK) cells** | CD14- CD3- CD123- CD66b- CD45RA+ CD56+ |
| **Mucosal-associated invariant T (MAIT) and NKT cells** | CD3+ CD66b- CD19- CD4- CD14- CD161+ TCR$\gamma\delta$- CD28+ CD16- |
| **B cells** | CD3- CD14- CD56- CD16- CD19+ CD20+ HLA-DR+ |
| **Plasmablasts** | CD3- CD14- CD16- CD66b- CD20- CD19+ CD56- CD38++ CD27++ |
| **Dendritic cells (DC)** |  |
| pDC | CD3- CD19- CD14- CD20- CD66b- HLA-DR+ CD11c- CD123+ |
| mDC | CD3- CD19- CD14- CD20- HLA-DR+ CD11c+ CD123- CD16- CD38+ CD294- |
| **Monocytes** | CD3- CD19- CD56- CD66b- CD14+ HLA-DR+ CD11c+ |
| *Classical* | CD14+ CD38+ |
| *Transitional* | CD14<sup>low</sup> CD38<sup>low</sup> |
| *Non-classical* | CD14- CD38- |
| **Neutrophils** | CD66b+ CD16+ HLA-DR- |

**Supplementary Table 3.** List of genes used to calculate zScores for each gene signature. Type I IFN, type II IFN, TNF-α and IL-1β signatures based on the study by Urrutia et al. (Cell Reports, 2016). IL-18 and IL-6 signatures based on Wikipathway. NFκB signature based on the Nanostring Immunology panel annotation file.

|  |  |
| --- | --- |
| **Type II IFN (IFN-γ)** | *CDKN1A, CXCL9, HLA-DMB, HLA-DPA1, HLA-DPB1, HLA-DRA, IDO1, JAK2, RARRES3, SLAMF7, SOCS1* |
| **TNF-α** | *C3, CCL4, CD44, CD83, IRAK2, NFKB2, NFKBIA, RELB, SOCS3, SRC, TNFAIP3* |
| **IL-1β** | *CCL2, CCL20, CXCL2, IL1A, IL1B, IL6, LILRB1, NFKB1, NFKBIZ, POU2F2* |
| **IL-18** | *ARG1, B2M, BAX, BCL2, BID, CASP3, CASP8, CCL18, CCL19, CCL2, CCL20, CCL3, CCL4, CCL5, CD36, CD81, CD83, CEBPB, CHUK, CTNNB1, CXCL2, FADD, FAS, FN1, ICAM1, IFNG, IKBKB, IL10, IL12B, IL13, IL18, IL18R1, IL18RAP, IL1B, IL2RA, IL6, IL9, IRAK1, IRF1, ITGA2B, LCK, MAPK1, MYD88, NFKB1, NFKB2, NFKBIA, NFKBIZ, NOS2, PRKCD, PTGS2, RELA, SOCS3, SPP1, TBX21, TNF, TNFAIP3, TNFSF11, TP53, TRAF1, TRAF6* |
| **IL-6** | *CEBP, HRAS, IL6, IL6R, IL6ST, JAK1, JAK2, JAK3, RAF1, STAT3* |
| **NFκB** | *CHUK, FADD, IKBKB, IKBKG, IL1A, IL1R1, MYD88, NFKB1, NFKBIA, RELA, TNF, TNFAIP3, TNFRSF1B, TRAF6* |

**Supplementary Table 4.** Top down-regulated and up-regulated gene in VEXAS patients and VEXAS-like patients compared to healthy controls.

| **Top down-regulated gene in VEXAS vs healthy controls comparison (q-value <0.05)** |  |  |
| --- | --- | --- |
| **Genes** | **estimate(log2FC)** | **q.value** |
| *CCR2* | -1,29864053 | 0,000694067 |
| *CSF1R* | -1,25732058 | 5,20926E-06 |
| *CMKLR1* | -1,165419347 | 0,000449572 |
| *CIITA* | -1,118451901 | 0,001136799 |
| *MSR1* | -1,089445902 | 0,002968737 |
| *CX3CR1* | -1,035615302 | 8,68107E-06 |
| *CASP10* | -1,033193025 | 0,002822755 |
| *CD86* | -0,948948417 | 0,00129542 |
| *CISH* | -0,938174083 | 0,001224332 |
| **Top up-regulated gene in VEXAS vs healthy controls comparison (q-value <0.05)** |  |  |
| **Genes** | **estimate(log2FC)** | **q.value** |
| *PLAU* | 1,49165211 | 8,11703E-06 |
| *NFKBIA* | 1,180562775 | 2,08163E-08 |
| *LTF* | 1,166018448 | 0,000618003 |
| *TNFAIP3* | 1,083489752 | 6,89703E-09 |
| *CTSG* | 1,07873244 | 0,00152962 |
| *CEACAM6* | 0,960270675 | 0,002327734 |
| *CXCL2* | 0,913118914 | 0,001319115 |
| *CEACAM8* | 0,906700247 | 0,003570479 |
| *CD83* | 0,869234642 | 8,11703E-06 |
| *CCL20* | 0,862340578 | 0,00110127 |

**Supplementary Table 5.** Down-regulated and up-regulated genes common in VEXAS and VEXAS-like in comparison to healthy controls.

|  |
| --- |
| **Down-regulated genes common in VEXAS and VEXAS-like in comparison to healthy controls (q-value <0.05)** |
| *CX3CR1, CISH, CXCR6, RORC, EOMES, KLRG1, TNFRSF8, IL4, IL7R, CD45RB, CD244, CD27, TP53, LCK, IL16, MBP, IKBKAP, IKBKE, S1PR1, IKZF1, NOS2, ICAM2, IKBKB, NFATC3* |
| **Up-regulated genes common in VEXAS and VEXAS-like in comparison to healthy controls (q-value <0.05)** |
| *PLAU, NFKBIA, TNFAIP3, CD83, NFKBIZ, ICAM1, RELB, CTNNB1, TRAF6, PSMD7* |

